# Physicians’ Knowledge and Experiences of Counseling on Complementary, Alternative, and Integrative Medicine in Cancer Care: A Qualitative Systematic Review

**DOI:** 10.1101/2025.02.26.25322744

**Authors:** Jeremy Y. Ng, Aimun Qadeer Shah, Lana Abu Narr, Alyssa Qian, Holger Cramer

**Affiliations:** Institute of General Practice and Interprofessional Care, University Hospital Tübingen, Tübingen, Germany; Robert Bosch Center for Integrative Medicine and Health, Bosch Health Campus, Stuttgart, Germany

**Keywords:** complementary and alternative medicine, integrative medicine, mindbody therapies, conventional medicine, barriers, perceptions

## Abstract

**Background:** Despite the increasing use of complementary, alternative, and integrative medicine (CAIM) use among cancer patients, there remains ambiguity surrounding physicians’ understanding and counseling practices in this area. The objective of this qualitative systematic review was to identify the knowledge and experiences of physicians who counsel patients on CAIM in the context of cancer.

**Methods:** MEDLINE, EMBASE, and AMED were systematically searched from inception to May 27, 2023. Reference lists of relevant review articles were also hand-searched. Eligible articles contained qualitative data focused on physicians’ perceived knowledge and experience pertaining to CAIM counseling for cancer care. Relevant findings were extracted and analyzed using a narrative synthesis approach to identify key themes and subthemes.

**Results:** Thirty-five articles were included (30 from database searching and 5 from hand-searching reference lists). Four main themes were identified: lack of knowledge and formal training on CAIM; distrust and concern about CAIM safety and/or efficacy; accepting CAIM as an important part of cancer care; and the communication dynamics between patients and physicians.

**Conclusions:** This study highlights that physicians recognize CAIM as an important component of person-centered and holistic cancer care; however, they have concerns about the safety and/or efficacy of these therapies. Accordingly, there is a need for improved education for physicians on the safety and effectiveness of CAIMs, to better equip them to effectively counsel patients in this area. Future research exploring the perspectives of medical trainees and other healthcare providers on CAIM for cancer is also warranted.

## 1. Introduction

Cancer is a complex, chronic health condition that is one of the leading causes of mortality globally. In 2022, an estimated 20 million new cancer cases were diagnosed and an approximate 9.7 million deaths were reported worldwide.^1^ The burden of cancer only continues to rise, and the incidence of all cancers combined is estimated to double by the year 2070.^2^ The conventional treatments for cancer include surgery, chemotherapy, and radiation; although these often cause various side effects such as nausea, fatigue, lack of motor coordination, and reduced quality of life.^3,4^ Accordingly, complementary, alternative, and integrative medicine (CAIM) have been popular among cancer patients in recent decades.

“Complementary medicines” are defined as non-conventional therapies used together with conventional therapies.^5,6^ “Alternative medicines” describe non-conventional therapies used in replacement of conventional therapies.^5,6^ “Integrative medicine” delivers conventional and non-conventional therapies together in a coordinated way among health professionals and institutions.^5,6^ CAIM encompasses a wide range of practices outside of mainstream medicine, including herbal remedies, acupuncture, yoga, meditation, and nutritional supplements.^7^

Studies have shown that CAIM use among cancer patients is highly prevalent, ranging from 25% to 51% in the last few decades,^8–11^ with some studies reporting even greater prevalence rates of CAIM use up to 87%.^12,13^ Patients use CAIM for a variety of reasons, including to address unmet needs, to mitigate side effects of conventional therapy, to improve quality of life,^8,12,14,15^ for general well-being,^8,12,16^ and to treat cancer or prevent it from spreading.^12^ However, the latter is very rare; less than 5% of cancer survivors who use CAIM actually do so to treat their cancer.^17^

Despite the increasing use of CAIM among cancer patients,^9,18^ the knowledge and experiences of physicians who counsel patients on CAIM remain unclear. Evidence suggests that most physicians feel they have limited knowledge of CAIM and are not up to date with the best evidence.^19,20^ Many physicians also do not feel competent to monitor patients’ use of CAIMs.^20^ The lack of knowledge and discomfort in monitoring CAIM usage contributes to providers’ reluctance to initiate discussions about CAIM with patients. For example, a literature review found that most general healthcare professionals and oncology experts were only likely to discuss CAIM use for cancer if their patient mentioned it first.^21^ On the other hand, patients are often hesitant to discuss their use of CAIM with providers for reasons including that their physician did not ask, feeling that their physician would disapprove, or believing that their physician was not interested in the topic.^11,15,22^

Physicians that fail to engage in effective discussions on CAIM could potentially impair communication and collaboration with their patients who use these therapies. This is especially concerning in the cancer care context where shared-decision making is important for treatment decisions and treatment adherence.^23,24^ Additionally, non-disclosed CAIM use poses a potential hazard to cancer patients as it may interfere with conventional treatments, with herb-drug interactions being a notable example.^25^ It is important for physicians to be interested in CAIM in order to address unmet information needs and to ensure the safety of their cancer patients.

Therefore, there is a need to evaluate the current state of the knowledge and experiences of physicians who counsel patients on the use of CAIM in cancer care. This qualitative systematic review synthesizes the available evidence to answer the question: What is the current knowledge and experiences of physicians who counsel patients on the use of CAIM in cancer care? Understanding physicians’ current knowledge is essential to identify potential barriers and facilitators for the integration of CAIM into cancer care.

## 2. Methods

### 2.1. Approach

A qualitative systematic review was conducted in accordance with the Preferred Reporting Items for Systematic Reviews and Meta-Analyses (PRISMA) guidelines.^26^ A systematic review of qualitative studies synthesizes findings from individual studies to capture the common experiences, perspectives, and attitudes of participants across multiple studies.^27^ Given the limited state of evidence regarding the range of physician experiences with CAIM in the context of cancer care, such an approach is particularly useful. A protocol was registered on PROSPERO (ID: CRD42023442324) on July 15, 2023. Study data were uploaded on Open Science Framework (https://doi.org/10.17605/OSF.IO/UQ39R), including the voting records for title and abstract screening (https://osf.io/ybdmg), full-text screening (https://osf.io/ac35f), Critical Appraisal Skills Programme (CASP) ratings (https://osf.io/pb8js), and the process work for the thematic analysis (https://osf.io/5p4jk).

### 2.2. Eligibility Criteria

The eligibility criteria were based on the Sample, Phenomenon of interest, Design, Evaluation, and Research type (SPIDER) framework.^28^ Eligible samples included physicians. The phenomena of interest were the knowledge and/or experiences of physicians when counseling on CAIM for cancer care. Qualifying CAIM therapies were assessed based on a broad operational definition of CAIM developed by Ng et al.,^29^ which was later adopted by the Cochrane Complementary Medicine group.^7^ However, “radiation therapy,” which is part of this all-inclusive list, was deliberately excluded for our purposes as it is known to be a conventional treatment for cancer. Eligible study designs were qualitative (e.g., interviews, focus groups, and observational studies) or mixed-method studies with a qualitative component (e.g., surveys). Studies with solely quantitative data were ineligible. The outcomes sought were qualitative data pertaining to physicians’ knowledge and experiences when counseling on CAIM in cancer care.

The research type included qualitative data or mixed-methods studies that generated qualitative data.

### 2.3. Searching and Screening

A literature search was conducted on May 27, 2023, with no year restrictions using three databases: MEDLINE, EMBASE, AMED. The search strategy included a comprehensive search string of CAIM terms^30^ which encompass 604 distinct therapies previously described in an operational definition of CAIM.^29^ The search string for CAIM terms was combined with various combinations of search terms for each of the following constructs: “physicians or oncologists,” “cancer,” and “qualitative or mixed method research.” The search strategy was adapted for each database using tailored subject headings and keywords. The complete search strategies for all three databases can be found in **Supplementary File 1**.

Endnote referencing software version 21^31^ was used to delete duplicates prior to screening. Rayyan systematic review software^32^ was used to screen all articles. AQ and LN first screened 50 articles for preliminary title and abstract screening, and 15 for preliminary full text screening. JYN, AS, AQ, and LN met to discuss pilot screenings and resolve selection differences. AQ and LN then independently screened the remaining titles and abstracts, and then full texts in duplicate. All disagreements between reviewers were resolved in consultation with AS, and then JYN if a resolution could not be reached.

Hand-searching of reference lists of relevant review articles was completed independently and in duplicate by AQ and LN. Relevant reviews were identified during screening and broadly examined the perspectives of healthcare providers on CAIM for cancer care. Articles identified as eligible by both reviewers were included, and discrepancies were resolved in a meeting with JYN, AS, AQ, and LN.

### 2.4. Data Extraction

A data extraction form was created *a priori* (JYN, AS, HC) to collect the following for each study: author, year, title, country, study objective, qualitative methodologies, theoretical underpinnings, sample characteristics, themes, main findings, challenges, limitations, and conclusions. A pilot extraction of five included articles was completed independently (LN, AQ, AS) and a meeting was held to discuss this to ensure consistency. LN and AQ then independently, and in duplicate, extracted data from the remaining articles. All extractions were reviewed by a third reviewer (JYN, AS, HC).

### 2.5. Quality Assessment

The quality of included studies was assessed using the CASP tool for qualitative studies^33^. This checklist includes ten items, with the first nine scored with a ‘yes,’ ‘no,’ or ‘can’t tell.’ The last question is an open-ended response; however, to ensure consistency across raters, we split this question based on the three ‘hints’ provided, and scored these three items with a ‘yes, ‘no,’ or ‘can’t tell,’ and each study could obtain a maximum score of 12 ‘yes’s.’ AQ and LN, independently and in duplicate, completed a pilot assessment of five articles, which was then discussed with AS to ensure consistency. Then, AQ and LN proceeded to independently and in duplicate, appraise the remaining articles.

### 2.6. Thematic Analysis

Following data extraction, the data was synthesized using a narrative synthesis approach^34^ involving the construction of descriptive themes and sub-themes. AQ and LN independently coded all included studies to synthesize common themes across in an iterative process. All authors then collaborated to discuss, revise, and finalize the themes (AQ, LN, AS, JYN, HC).

## 3. Results

### 3.1. Search Results

A total of 4344 items were retrieved across the database searches, of which 3279 were unique. 3062 titles and abstracts were eliminated, leaving 212 that underwent full text screening. Upon screening by full-text, 182 were deemed ineligible, leaving 30 eligible articles. The database searches also revealed six relevant review articles which were hand-searched systematically to identify an additional five eligible articles. Thus, a total of 35 articles are included in this systematic review^35–69^ as seen in **Figure 1**.

**Figure 1:**
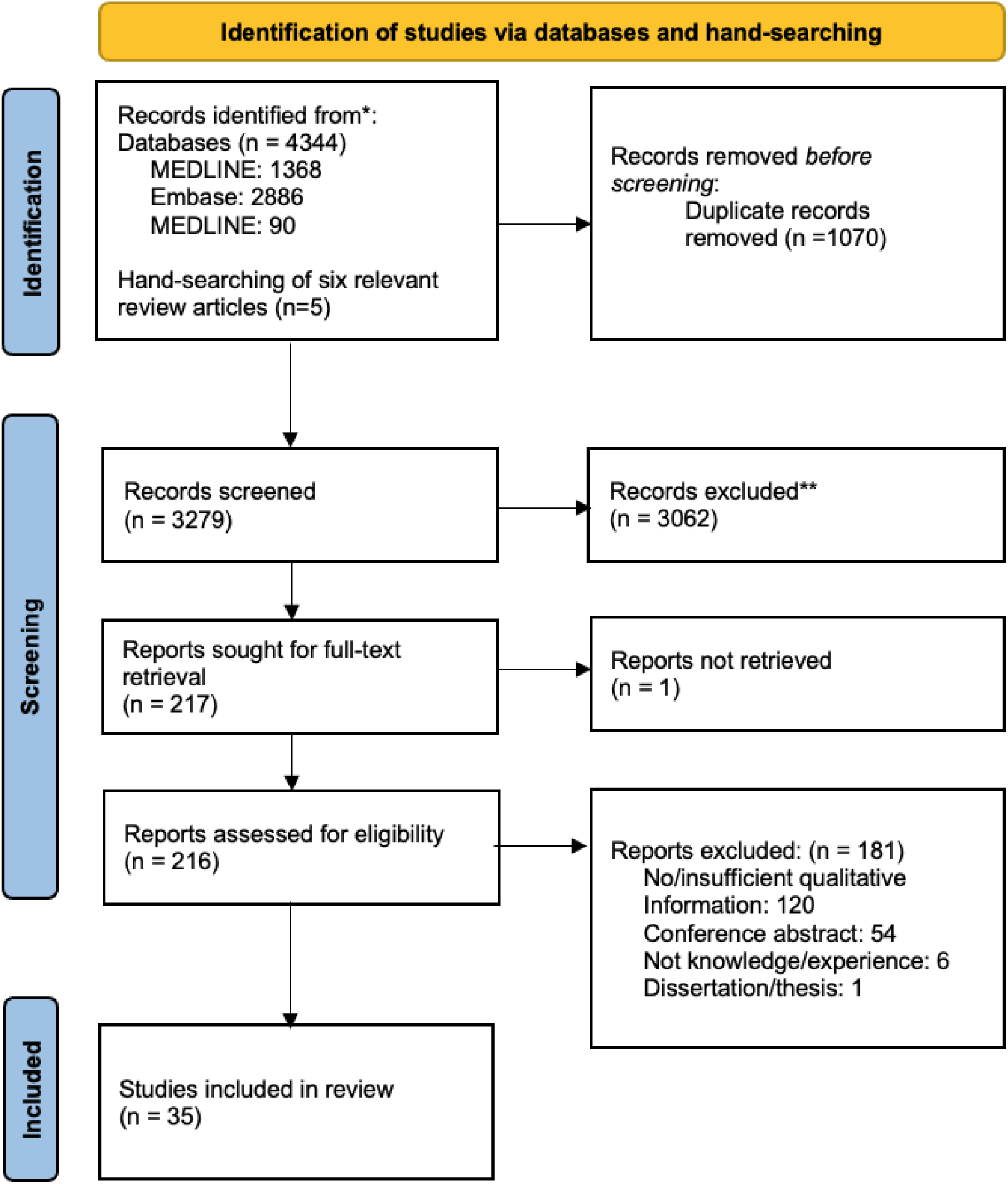
PRISMA Flow Chart of Study Selection Process26

### 3.2. Characteristics of Included Studies

As shown in **Supplementary File 1** and **Supplementary File 2**, eligible articles were published between 1996 and 2023. The largest number of studies originated from Canada (n=11), followed by Germany (n=7) and Australia (n=4). The studies predominantly focused on the knowledge and experiences of physicians counseling about CAIM in general (n=21), although some focused on lifestyle (diet and exercise) therapies (n=10), cannabis (n=2), self-help groups (n=1), and non-pharmacological interventions (n=1). The most common research method used were qualitative interviews (n=25). Nineteen studies focused exclusively on physicians, while 16 studies included multiple health professionals and the extracted findings were limited only to physicians. Most physicians were oncologists (n=19) and general/family practitioners (n=12), but other specialties were also represented such as hematologists, palliative care physicians, and internal medicine physicians.

### 3.3 Results from Quality Assessment

All 35 articles were included in the review regardless of their quality ratings, presented in **Table 1**. Three studies scored 11/12 possible ‘yes’ responses, 9 studies scored 10/12 possible ‘yes’ responses, 9 studies scored 9/12 possible ‘yes’ responses, 7 studies scored 8/12 possible ‘yes’ responses, 6 studies scored 7/12 ‘yes’ responses, and 1 study scored 6/12 ‘yes’ responses. Common potential sources of bias were that included studies were not clear about the relationship between the researcher and the participants, and they did not discuss whether or how their findings could be transferred to other populations or considered in other research.

**Table 1:** Critical Appraisal Skills Programme (CASP) Qualitative Research Checklist Quality Appraisal of Included Studies (N=35)

| Title | Author | Was there a clear statement of the aims of the research? | Is a qualitative methodology appropriate? | Was the research design appropriate to address the aims of the research? | Was the recruitment strategy appropriate to the aims of the research? | Was the data collected in a way that addressed the research issue? | Has the relationship between researcher and participants been adequately considered? | Have ethical issues been taken into consideration? | Was the data analysis sufficiently rigorous? | Is there a clear statement of findings? | Does researcher discuss the contribution the study makes to existing knowledge or understanding? | Do they identify new areas where research is necessary? | Do they discuss whether or how the findings can be transferred to other populations or considered other ways the research may be used? | Total |
| --- | --- | --- | --- | --- | --- | --- | --- | --- | --- | --- | --- | --- | --- | --- |
| Are We Meeting Cancer Patient Needs? Complementary and Alternative Medicine Use Among Saudi Cancer Patients: A Qualitative Study of Patients and Healthcare Professionals' Views | Alqahtani et al., 2018<sup>35</sup> | Yes | Yes | Yes | Can't tell | Yes | No | Yes | Yes | Yes | Yes | Yes | Yes | Yes: 10<br>No: 1<br>Can't tell: 1 |
| Patient and Medical Oncologists' Perspectives on Prescribed Lifestyle Intervention—Experiences of Women with Breast Cancer and Providers | Balneaves et al., 2020<sup>36</sup> | Yes | Yes | Yes | Yes | Yes | No | Yes | Yes | Yes | Yes | Yes | Yes | Yes: 11<br>No: 1<br>Can't tell: 0 |
| "Part of the Conversation": A Qualitative | Balneaves & Watling, 2022<sup>37</sup> | Yes | Yes | Yes | Can't tell | Yes | No | Yes | No | Yes | Yes | Yes | No | Yes: 8<br>No: 3<br>Can't tell: 1 |
| Study of Oncology Healthcare Professionals' Experiences of Integrating Standardized Assessment and Documentation of Complementary Medicine |  |  |  |  |  |  |  |  |  |  |  |  |  |  |
| Supportive Care and Osteopathic Medicine in Pediatric Oncology: Perspectives of Current Oncology Clinicians, Caregivers, and Patients | Belsky et al., 2021<sup>38</sup> | No | Yes | No | Yes | Yes | Yes | Yes | Yes | Yes | Yes | Yes | No | Yes: 9<br>No: 3<br>Can't tell: 0 |
| Communication and Integration: A Qualitative Analysis of Perspectives Among Middle Eastern Oncology Healthcare Professionals on the Integration of Complementary Medicine in Supportive Cancer Care | Ben-Arye et al., 2015<sup>39</sup> | Yes | Yes | No | No | No | No | Yes | No | Yes | Yes | Yes | Yes | Yes: 7<br>No: 5<br>Can't tell: 0 |
| Physicians' Attitudes Toward Patients' Use of Alternative Cancer Therapies | Bourgeault, 1996<sup>40</sup> | Yes | Yes | Yes | Yes | Yes | No | No | Yes | Yes | Yes | No | Yes | Yes: 9<br>No: 3<br>Can't tell: 0 |
| Oncology Clinicians' Accounts of Discussing Complementary and Alternative Medicine With Their Patients | Broom & Adams, 2009<sup>41</sup> | Yes | Yes | Yes | Yes | Yes | No | Yes | Yes | Yes | Yes | No | No | Yes: 9<br>No: 3<br>Can't tell: 0 |
| Traditional Medicines, Collective | Broom & Doron, 2012<sup>42</sup> | No | Yes | Yes | Yes | Yes | No | Yes | Yes | Yes | Yes | Yes | Yes | Yes: 10<br>No: 2<br>Can't tell: 0 |
| Negotiation, and Representations of Risk in Indian Cancer Care |  |  |  |  |  |  |  |  |  |  |  |  |  |  |
| Bridging the Gap Between Attitudes and Action: A Qualitative Exploration of Clinician and Exercise Professional's Perceptions to Increase Opportunities for Exercise Counselling and Referral in Cancer Care | Caperchione et al., 2022<sup>43</sup> | Yes | Yes | Yes | Yes | Yes | No | Yes | Yes | Yes | Yes | Yes | No | Yes: 10<br>No: 2<br>Can't tell: 0 |
| Changing Physicians' Attitudes Toward Self-help Groups: An Educational Intervention | Carroll et al., 2000<sup>44</sup> | Yes | Yes | Yes | Yes | Yes | No | No | Yes | Yes | Yes | Yes | No | Yes: 9<br>No: 3<br>Can't tell: 0 |
| Oncologists' Experiences of Discussing Complementary and Alternative Treatment Options With Their Cancer Patients. A Qualitative Analysis | Corina et al., 2016<sup>45</sup> | Yes | Yes | Yes | No | No | No | Yes | No | Yes | Yes | No | No | Yes: 6<br>No: 6<br>Can't tell: 0 |
| Complementary Medicine for Cancer Patients in General Practice: Qualitative Interviews with German General Practitioners | Dahlhaus et al., 2015<sup>46</sup> | Yes | Yes | Yes | Yes | Yes | No | Yes | Yes | Yes | Yes | No | No | Yes: 9<br>No: 3<br>Can't tell: 0 |
| A Qualitative Study of Patient and Healthcare Provider Perspectives on Building Multiphasic Exercise Prehabilitation | Daun et al., 2022<sup>47</sup> | Yes | Yes | Yes | Can't tell | Yes | Can't tell | Yes | Yes | Yes | Yes | Yes | Yes | Yes: 10<br>No: 0<br>Can't tell: 2 |
| into the Surgical Care Pathway for Head and Neck Cancer |  |  |  |  |  |  |  |  |  |  |  |  |  |  |
| The Challenge of Timing: A Qualitative Study on Clinician and Patient Perspectives About Implementing Exercise-Based Rehabilitation in an Acute Cancer Treatment Setting | Dennett et al., 2020<sup>48</sup> | Yes | Yes | Yes | Yes | Yes | Can't tell | Yes | Yes | Yes | Yes | Yes | No | Yes: 10<br>No: 1<br>Can't tell: 1 |
| A Comparison of Physician and Patient Perspectives on Unconventional Cancer Therapies | Gray et al., 1997<sup>50</sup> | Yes | Yes | Yes | Can't tell | Yes | Can't tell | No | Yes | Yes | Yes | Yes | No | Yes: 8<br>No: 2<br>Can't tell: 2 |
| Physician Perspectives on Unconventional Cancer Therapies | Gray et al., 1998<sup>49</sup> | No | Yes | Yes | Can't tell | Yes | Yes | No | Yes | Yes | No | No | Yes | Yes: 7<br>No: 4<br>Can't tell: 1 |
| What Hinders Healthcare Professionals in Promoting Physical Activity Towards Cancer Patients? The Influencing Role of Healthcare Professionals' Concerns, Perceived Patient Characteristics and Perceived Structural Factors? | Haussmann et al., 2018<sup>51</sup> | Yes | Yes | Can't tell | Yes | Yes | Yes | Yes | Yes | Yes | Yes | Yes | Yes | Yes: 11<br>No: 0<br>Can't tell: 1 |
| On Caring and Sharing—Addressing Psychological, Biographical, and Spiritual Aspects in Integrative Cancer Care: A | Kienle et al 2018a<sup>52</sup> | Yes | Yes | Yes | No | Yes | Yes | Yes | Yes | Yes | Yes | Yes | No | Yes: 10<br>No: 2<br>Can't tell: 0 |
| Qualitative Interview Study on Physicians' Perspectives |  |  |  |  |  |  |  |  |  |  |  |  |  |  |
| The Subjective Dimension of Integrative Cancer Care: A Qualitative Study Exploring the Perspectives, Themes, and Observations of Experienced Doctors from the Area of Anthroposophic Medicine | Kienle et al 2018b<sup>53</sup> | Yes | Yes | Yes | No | Yes | No | Yes | Yes | Yes | Yes | Yes | No | Yes: 9<br>No: 3<br>Can't tell: 0 |
| German Physicians' Perceptions and Views on Complementary Medicine in Pediatric Oncology: A Qualitative Study | Klatt et al., 2023<sup>54</sup> | Yes | Yes | Yes | No | Yes | No | Yes | Yes | Yes | Yes | No | No | Yes: 8<br>No: 4<br>Can't tell: 1 |
| Lifestyle Advice to Cancer Survivors: A Qualitative Study on the Perspectives of Health Professionals | Koutoukidis et al., 2017<sup>69</sup> | Yes | Yes | Yes | No | Yes | Yes | Yes | Yes | Yes | Yes | Yes | Yes | Yes: 11<br>No: 1<br>Can't tell: 0 |
| Academic Doctors' Views of Complementary and Alternative Medicine (CAM) and Its Role Within the NHS: An Exploratory Qualitative Study | Maha & Shaw, 2007<sup>55</sup> | Yes | Yes | Yes | No | Yes | No | No | Yes | Yes | Yes | Yes | Yes | Yes: 9<br>No: 3<br>Can't tell: 0 |
| Yoga in Adult Cancer: A Pilot Survey of Attitudes and Beliefs Among Oncologists | McCall & Heneghan, 2015<sup>56</sup> | Yes | Yes | No | No | No | Yes | Yes | No | Yes | Yes | Yes | Yes | Yes: 8<br>No: 4<br>Can't tell: 0 |
| How is Complementary Medicine | Mentink et al, 2022<sup>57</sup> | Yes | Yes | Yes | Yes | Yes | Yes | Yes | No | Yes | Yes | Yes | No | Yes: 10<br>No: 2<br>Can't tell: 0 |
| Discussed in Oncology? Observing Real-Life Communication Between Clinicians and Patients with Advanced Cancer |  |  |  |  |  |  |  |  |  |  |  |  |  |  |
| Oncologists' and Naturopaths' Nutrition Beliefs and Practices | Novak & Chapman, 2001<sup>58</sup> | Yes | Yes | Yes | Yes | Yes | Yes | Yes | Yes | Yes | No | No | No | Yes: 9<br>No: 3<br>Can't tell: 0 |
| Complementary Therapy Use by Cancer Patients: Physicians' Perceptions, Attitudes, and Ideas | O'Beirne et al., 2004<sup>59</sup> | Yes | Yes | Yes | Yes | Yes | No | Yes | No | Yes | No | No | No | Yes: 7<br>No: 5<br>Can't tell: 0 |
| Perspectives of Pediatric Oncologists and Palliative Care Physicians on the Therapeutic Use of Cannabis in Children with Cancer | Oberoi et al., 2022<sup>60</sup> | Yes | No | No | Yes | No | Yes | Yes | Can't tell | Yes | Yes | Yes | No | Yes: 7<br>No: 4<br>Can't tell: 1 |
| Convergent Priorities and Tensions: a Qualitative Study of the Integration of Complementary and Alternative Therapies with Conventional Cancer Treatment | River et al., 2018<sup>61</sup> | Yes | Yes | Yes | Yes | Yes | No | Yes | Yes | Yes | Yes | Yes | No | Yes: 10<br>No: 2<br>Can't tell: 0 |
| Mind the Gap! Lay and Medical Perceptions of Risks Associated with the Use of Alternative Treatment and Conventional Medicine | Salamonsen, 2015<sup>62</sup> | Yes | Yes | Yes | Yes | Yes | No | Yes | Yes | Yes | Yes | Yes | No | Yes: 10<br>No: 2<br>Can't tell: 0 |
| 'They Don't Ask Me So I Don't Tell Them': | Shelley et al., 2009<sup>63</sup> | Yes | Yes | Yes | Yes | No | No | No | No | Yes | Yes | Yes | Yes | Yes: 8<br>No: 4<br>Can't tell: 0 |
| Patient-Clinician Communication About Traditional, Complementary, and Alternative Medicine |  |  |  |  |  |  |  |  |  |  |  |  |  |  |
| Attitudes Toward Complementary and Alternative Medicine Amongst Oncology Professionals in Brazil | Siegel et al., 2016<sup>64</sup> | No | Yes | Yes | Yes | Yes | No | Yes | Yes | Yes | Yes | No | No | Yes: 8<br>No: 4<br>Can't tell: 0 |
| Considerations in Developing and Delivering a Non-Pharmacological Intervention for Symptom Management in Lung Cancer: The Views of Healthcare Professionals | Wagland et al., 2012<sup>65</sup> | Yes | Yes | Yes | Can't tell | Yes | No | Yes | Yes | Yes | No | No | No | Yes: 7<br>No: 4<br>Can't tell: 1 |
| 'Probably Better Than Any Medication We Can Give You' | Waterland et al., 2020<sup>66</sup> | Yes | Yes | Yes | No | Yes | Yes | Yes | Yes | Yes | No | Yes | No | Yes: 9<br>No: 3<br>Can't tell: 0 |
| Communication About Complementary and Alternative Medicine When Patients Decline Conventional Cancer Treatment: Patients' and Physicians' Experiences | Wode et al., 2023<sup>67</sup> | Yes | Yes | Yes | Yes | No | No | Yes | Yes | Yes | Yes | No | No | Yes: 8<br>No: 4<br>Can't tell: 0 |
| Medical cannabis: An Oxymoron? Physicians' Perceptions of Medical Cannabis | Zolotov et al., 2018<sup>68</sup> | Yes | Yes | Yes | No | No | No | Yes | Can't tell | Yes | Yes | Yes | No | Yes: 7<br>No: 4<br>Can't tell: 1 |

### 3.4. Findings From Thematic Analysis

Four main themes were identified from our analysis and are described below. Representative quotes for each theme and sub-theme are displayed in **Table 2**.

**Table 2:** Themes, Subthemes, and Representative Quotes from Included Qualitative Studies on Physicians’ Knowledge and Experiences Counselling on Complementary, Alternative, and Integrative Medicine (CAIM) in Cancer Care.

| **Theme #1: Lack of CAIM Knowledge & Formal Training** |  |
| --- | --- |
| **Sub-themes** | **Representative Quotes** |
| **Physician lack of confidence in CAIM counseling** | "Clinicians reported having low awareness and lacked confidence about what to advocate in terms of rehabilitation and exercise, resulting in vague advice being given to patients."; "There was also poor knowledge about what resources were available for rehabilitation."<sup>48</sup><br>[It's] as important as all the pharmacological treatment but probably as GPs we don't do it enough and don't have enough confidence in giving recommendations regarding exercise and nutrition."; "I don't think I'm qualified to give very specific dietary advice."<sup>66</sup> |
| **Need for credible and formal sources of CAIM information** | "Should there be education ... yes, but realistically who's going to do it?"; "I mean there probably should be more information for us on CAM."; "There isn't a lot of education-based sessions ..."<sup>41</sup><br>"I think that's the real issue; I don't think I have a real resource, I'm just using my brain, my common sense. I don't think I've ever had any tuition about diet and cancer."<sup>66</sup><br>"I think having resources that are simple and easy to use that are fairly generic so that [they] can be used for most cancers and a handout for patients would be incredibly useful."<sup>66</sup> |
| **CAIM being outside of physicians' scope of practice** | "Our priority here is to develop our medical service through a purely medical approach, not by something like CAM"; I am a doctor of medicine, not a CAM specialist.<sup>35</sup><br>"These physicians did not want to seem entirely dismissive of patients' interests but nevertheless did not wish to spend their time discussing unconventional therapies, either because they did not think they were worth discussing or because they saw such approaches as outside their expertise."<sup>50</sup> |
| **Theme #2: Distrust and Concern about CAIM Safety and/or Efficacy** |  |
| **Sub-themes** | **Representative Quotes** |
| **Concerns for patient wellbeing (health and financial consequences)** | "And for me, it was a big concern around toxicity and potential harm, so I thought that encouraging the conversation and disclosing could help, hopefully, patient safety and increase awareness of the products [used]."<sup>37</sup> |
| **Lack of rigorous research/evidence-based findings** | "I don't think CTM should be integrated into standard oncology care unless controlled, double-blind phase 3 trials are conducted which prove the benefits for patients."<sup>39</sup> |
| **Preference for conventional medicine** | "We need to stick with conventional medicine."<sup>35</sup><br>"An alternative therapy might not be harmful per se, but if [it] deprives or delays a patient from receiving curative therapy, . . . then [it is] harmful."<sup>40</sup> |
| **Concerns with misleading patients** | "My main objection to [cannabis] use in palliative care is that families typically wish to start it as a curative, not palliative adjunct, treatment based on certain misrepresentations online about cannabis as miracle cure."<sup>60</sup><br>"In addition, some of the physicians felt that alternative cancer therapies are harmful psychologically, because of the false hope they give patients, and financially, because of their often exorbitant cost."<sup>40</sup> |
| **Theme #3: Accepting CAIM as an Important Part of Cancer Care** |  |
| **Sub-themes** | **Representative Quotes** |
| **CAIM is clinically relevant in cancer care** | "Several respondents envisioned the integration of CTM, primarily within supportive and palliative cancer care clinics."<sup>39</sup> |
|  | "Oncologists and most patients believed the intervention should be offered as a standard part of breast cancer care, and as early as possible within the cancer trajectory..."<sup>36</sup> |
| **CAIM acceptance in the context of holistic cancer care** | "An essential task for me is to strengthen patients in dealing with their disease, in learning how to tackle the disease with their whole personality."<sup>52,53</sup><br>"I see it as being a very good thing when I say that we also provide aromatherapy. That is something that patients like very much indeed."<sup>45</sup> |
| **Recognition of the importance of interdisciplinary collaboration (with CAIM providers)** | "Other respondents emphasized the need for good communication and close collaboration between oncologists and CTM practitioners."<sup>39</sup><br>"Those who have mastered alternative medicine should join the medical doctors at the clinics... This must be accomplished through joint work of the physicians and the alternative medicine practitioners."<sup>39</sup> |
| **Theme #4: Patient-Physician Communication Dynamic** |  |
| **Sub-themes** | **Representative Quotes** |
| **Patient-related Barriers** | "Physicians thought that most of their patients do not disclose their use or contemplation of use of complementary therapies. One participant who accepts referrals from other physicians to counsel patients on complementary therapy believed that only 60% of his own patients told him about their use of complementary therapies."<sup>65</sup> |
| **Physician-related Barriers** | "However, they typically avoid the topic where possible and do not see discussion of unconventional therapies as a priority use of their time."<sup>49, 50</sup> |
| **Clinical administration- and policy-related Barriers** | "Ten-minute consultations are simply spinning the wheels in the mud. You can't do anything, because you don't have time to do anything."<sup>66</sup><br>"I don't have enough time to do it all [provision of lifestyle advice] properly and sensitively."<sup>69</sup> |
| **The role of shared decision-making and patient-centered care in CAIM counseling** | "We felt that we weren't really catering for our patients appropriately in that sort of holistic fashion, so we've wanted them to be able to use music therapy, acupuncture, herbal medicine advice, massage, whatever, but do it in a way that is controlled and evidence-based."<sup>61</sup> |

#### Theme 1: Lack of CAIM Knowledge & Formal Training

In 28 out of the 35 included studies, physicians suggested a lack of CAIM knowledge and formal training.^35,37–41,43,45,46,48–51,53–61,63,64,66–69^ Within this theme, the following sub- themes were identified: physician lack of confidence CAIM counseling, need for credible and formal sources of CAIM information, and CAIM being outside of physician’s scope of practice.

##### Sub-theme 1.1: Physicians’ Lack of Confidence in CAIM Counseling

The majority of studies concluded that physicians feel uncertain and skeptical about the topic of CAIM, and accordingly, lack confidence when counselling cancer patients on the subject. This uncertainty stems from physicians’ perceptions of insufficient knowledge on CAIM.^37,43,45,46,49,50,52–59,61,64,66–69^ Some physicians were also fearful that CAIM may interact negative and interfere with conventional medicine^41,57,67^; however, studies also highlighted that physicians have limited understanding on the potential side effects associated with CAIM.^37,54,67^ Additionally, several articles suggested that there is a shortage in training on CAIM consultation which prevents physicians from becoming more familiar with CAIM.37,39,40,43,51,59,63,64,66-69

***Sub-theme 1.2: Demand for Credible and Formal Sources of CAIM Information*** Physicians voiced the necessity for implementing standardized CAIM education and training across institutions.^37,39,41,43,46,50,66^ For additional effectiveness, the education should cover both practical and clinical aspects of CAIM consultation.^43^ One study recommended the integration of a concise introductory course into undergraduate medical curricula, suggesting that the education does not necessarily need to delve deep into specific CAIM therapies, but should be used as an approach to increase general awareness of CAIM through a brief overview of therapies.^55^ Furthermore, many studies concluded that physicians had difficulty accessing evidence based CAIM resources due to insufficient experimental data and a lack of credible sources.^37,40,46,48–50,54–57,60,66^ The wide range of CAIMs that exist made it even more arduous for physicians to locate sources on or learn more about the CAIMs used by their patients.^37^

##### Sub-theme 1.3: CAIM Being Outside of Physicians’ Scope of Practice

Many physicians saw CAIM as outside their scope of practice, stating that consulting about CAIM does not fit their clinical role.^35,37,45,46,49,54^ Physicians believed that assessing CAIM may disrupt their responsibilities in the healthcare team or mislead patients about the nature of their professional role.^37^ Physicians also suggested that conventional medicine practice should have priority over CAIM.^35,41,46,50,55^ Overall, many agreed that CAIM consultation is mutually exclusive to their domains of practice^35,45,46,49,54^ and that CAIM assessments should be completed by allied health professionals who are more familiar with the topic.^37,43,47,66^ Physicians also perceived that other types of healthcare providers could provide more detailed information on CAIM’s risks and benefits,^37,43^ safety and effectiveness,^43^ and patient instructions on CAIM usage than physicians.^47^

#### Theme 2: Distrust And Concern About CAIM Safety and/or Efficacy

In 24 out of the 35 included studies, physicians reported a distrust and concern about CAIM safety and/or efficacy.^35,37,39–42,45,46,48–50,54–64,67,68^ Within this theme, the following sub-themes were identified: concerns for patient wellbeing (health- and financial-related), lack of rigorous research or evidence-based findings, preference for conventional medicine, and concerns regarding misleading patients.

***Subtheme 2.1: Concerns For Patient Wellbeing (Health- And Financial-Related)***

Physicians across several studies expressed concern regarding the potential for harm that CAIM interventions may pose to patient well-being, encompassing both health and financial risks.^35,37,40–42,49,56,59–62,67^ Some physicians highlighted the risk of toxicity and physical harm associated with CAIM interventions due to the potential impurities in their compositions.^35,41,42^ Concurrently, there was concern that unqualified providers exploited CAIM for financial gain,^45,50,55^ leading to an unnecessary financial burden on patients seeking these therapies.^40,59–61,67^ Physicians’ skepticism translated into their conversations with patients, whereby they often emphasized the adverse physical and financial consequences of CAIM use in an attempt to dissuade patients from pursuing these interventions.^55,67^

##### Subtheme 2.2: Lack Of Rigorous Research and Evidence-Based Findings

Physicians expressed significant concerns regarding the lack of rigorous, evidence-based research such as controlled trials and safety assessments, when counselling patients on CAIM.^37,39–41,45,46,49,50,55–64,67,68^ For example, physicians in one article cited the absence of double-blinded phase three trials on CAIM as a primary reason for their skepticism regarding its integration into standard oncology care.^39^ The dearth of credible scientific evidence acted as a barrier to providing comprehensive advice or suggestions to patients, further complicating physician-patient decision-making regarding CAIM.49,50,63

##### Subtheme 2.3: Preference For Conventional Medicine

Several articles highlighted physicians’ inclination towards conventional medicine over CAIM.^35,39,41,48,50,60,61,64,68^ Physicians indicated their preference for conventional medicine, stating their beliefs that conventional medicine is superior to CAIM and hence, patients should “stick with conventional medicine.”^35,39,41^ Additionally, concerns were raised about the potential negative interactions between CAIM and conventional therapies, leading physicians to prioritize conventional treatments to avoid patient harm^40–42^ Limitations in health resources and the perceived “lack of capacity for new services” was another factor that compelled physicians to prioritize conventional therapies over CAIM interventions.^64^

##### Subtheme 2.4: Concerns With Misleading Patients

Finally, physicians worried about the psychological distress that CAIM use may cause to patients by promoting false promises of a cure or encouraging irrational and unrealistic expectations.^40,41,45,50,55,59,60^ This concern was especially pronounced in palliative care settings, where patients may perceive CAIM interventions as “a miracle cures” as opposed to adjunct therapies.^60^ Moreover, physicians were apprehensive about the misleading information proliferating online or through marketing efforts which inaccurately portrayed CAIM as more “natural” and therefore superior to conventional pharmacotherapies.^50,60^

#### Theme 3: Accepting CAIM as an Important Part of Cancer Care

In 18 of the 35 included articles, the theme of accepting CAIM as an important part of cancer care emerged.^37–40,42,43,45,49,50,52–56,64,66–68^ Three subthemes were identified: CAIM is clinically relevant in cancer care, CAIM acceptance in the context of holistic cancer care, and recognition of the importance of interdisciplinary collaboration with CAIM providers.

##### Subtheme 3.1: CAIM Is Clinically Relevant in Cancer Care

Physicians acknowledged that CAIM was clinically relevant in the context of cancer care, as reported by several articles.^36,38–40,45,49,50,52,53,56,64,66,68^ Some physicians viewed CAIM as a beneficial adjunct to standard treatment modalities.^36,38,40,49,50,56,64,66,68^ They felt it could offer additional options when conventional treatments failed to yield improvements in patients’ conditions.^38,40^ Physicians also recognized the potential of CAIM interventions such as lifestyle modifications (i.e., diet, yoga, exercise) to enhance tolerance of conventional treatments.^40,56,66^ Additionally, physicians identified CAIM interventions as valuable additions to patient care plans particularly in palliative care settings.^36,38,39^

##### Subtheme 3.2: CAIM Acceptance in the Context of Holistic Cancer Care

Physicians reported acceptance for CAIM due its facilitation of holistic, patient-centered, and higher quality care.^36,45,49,50,52,53,55,67^ CAIM interventions were recognized for their potential to manage side effects from standard treatment, thereby contributing to holistic health improvement.^36,38,55,68^ Physicians also emphasized the importance of addressing patients’ psychological wellbeing as part of holistic care, including their fear and attitudes towards life and death, and suggested that CAIM offered an avenue for such considerations.^52,53,67^ Furthermore, physicians recognized that the integration of CAIM in cancer care can empower patients to regain autonomy and control, fostering a patient-centered and shared-decision making approach.^49,50,52,53^ Accordingly, some physicians actively sought out additional resources outside of conventional treatments, demonstrating their interest in incorporating CAIM as an important component of cancer _care._38,39,52,53

##### Subtheme 3.3: The Importance of Interdisciplinary Collaboration with CAIM Providers

In some qualitative studies, physicians underscored the need to adopt a collaborative team approach and demonstrated a readiness to working alongside CAIM specialists.^39,45,54,66^ Physicians also emphasized the importance of improving communication with CAIM providers to ensure a collective and consistent understanding of patients’ treatment plans.^39^ For example, concerning exercise, physicians identified a need for direct collaboration with exercise physiologists.^43^ However, a major barrier to implementing a collaborative approach lay in the lack or complexity of existing referral pathways for directing patients to CAIM specialists.^43,45,66^

#### Theme 4: The Communication Dynamics Between Patients and Physicians

In 21 out of the 35 included studies, physicians discussed the patient-physician communication dynamic revolving CAIM.^36,37,39–42,45,47,49–55,59,61,63,65,66,69^ Within this theme, the following sub-themes were identified: patient-related barriers, physician-related barriers, clinical administration- and policy-related barriers, and the role of shared decision-making and patient-centered care in CAIM counseling.

##### Sub-theme 4.1: Patient-Related Barriers

The most prominent barrier as it concerns the patient-physician dynamic was patients’ lack of disclosure about CAIM usage.^37,39,42,59,65^ Physicians reported that patients often refrained from disclosing CAIM use due to their fear of being advised to stop,^37^ as well as to avoid physicians’ negative reactions such as anger,^39^ disinterest,^42^ reluctance to discuss it,^42^ and perceived lack of engagement during consultations.^65^ Physicians in one article suggested that patients chose not to disclose CAIM usage as they believed physicians could not provide helpful information.^42^ The lack of transparency on CAIM use posed several drawbacks, as physicians voiced being unable to caution patients about the potential for harm and toxicity associated with CAIM^37,39^ or being unable to educate and support patients in making informed decisions.^37^

##### Sub-theme 4.2: Physician-Related Barriers

Physician-related barriers centered on the belief that physicians often overlooked CAIM as a critical part of their roles.^49–51,55,63,69^ Physicians believed CAIM should not be considered a priority, particularly in the absence of robust scientific evidence.^55^ Additionally, physicians did not actively initiate discussions about CAIMs with patients; instead, CAIMs were only addressed if patients raised the topic.^55,63^

##### Sub-theme 4.3: Clinical Administration- and Policy-Related Barriers

Clinical policies and administrative procedures also posed a barrier to counselling about CAIM. The primary concern was the shortage of time during consultation sessions, which hindered discussions with patients regarding both conventional and CAIM approaches.^37,45,50,51,54,66,69^ Some physicians advocated for CAIM being included in standard care conversations and deserving of more attention in medical practices.^37,39^ Incorporating CAIM as part of the standard procedures would address several other barriers raised by physicians, including challenges in systematically assessing CAIM or identifying the exact CAIMs patients used.^37^ Additional regulatory barriers included insufficient funding to train staff on CAIM ^37^, limited clinic space to accommodate additional patients interested in CAIM,^65^ and the lack of established referral pathways to consult on CAIM use.^65^

##### Sub-theme 4.4: The Role of Shared Decision-Making and Patient-Centered Care in CAIM Counseling

Many studies underscored the importance of CAIM in shared decision-making and patient-centered care models.^36,37,39–41,45,47,49–53,59,61,63,66,69^ Physicians highlighted that patients’ use of CAIM could induce positive emotions and mindsets that may facilitate beneficial treatment decisions and aid in recovery.^36,40,41,49,50,59,61,63^ Moreover, physicians emphasized the importance of cultivating a trusting relationship with their patients to ensure full awareness about their CAIM usage, thereby enhancing the quality of care they provide.^45,47^ Overall, participants across the studies concurred that being open and accepting about patients’ utilization of CAIM boosted the physician-patient relationship.^37,45,47,49,50^ Conversely, misaligned views on CAIM could undermine such a dynamic.^49,50^ Hence, even if physicians held reservations about CAIM’s effectiveness, they respected patients’ autonomy in utilizing them^39,52,53,66^ and tolerated such actions as long as they did not reasonably interfere with their conventional treatment or overall wellbeing.^51,61^

## 4. Discussion

The objective of this qualitative systematic review was to synthesize the available evidence regarding the current knowledge and experiences of physicians who counsel patients on the use of CAIM in the context of cancer care. Through a comprehensive review of 35 eligible articles, four main themes were identified: lack of knowledge and formal training on CAIM; distrust and concern about CAIM safety and/or efficacy; accepting CAIM as an important part of cancer care; and communication dynamics between patients and physicians.

### 4.1. Comparative Literature

#### Lack of Knowledge and Formal Training on CAIM

Findings across the literature support that physicians’ knowledge and training related to CAIMs was limited or non-existent. For example, a recent systematic review found that physicians had poor knowledge of CAIM in cancer care, and that the lack of information on their safety and efficacy was a major barrier to discussing CAIM use with their patients.^20^ Similar to our review findings though, physicians in the literature are more familiar with mind-body therapies such as yoga as compared to biologically-based therapies such as herbal medications or vitamin supplements.^70,71^ This is especially concerning in the cancer care context as biologically based CAIMs are known to be the most popular among cancer patients,^22,72^ and their use is known to significantly increase after a cancer diagnosis.^73^

Interestingly, this review also found that while physicians desired more education about CAIM and believe it is important, they also perceived CAIM to be outside of their scope of practice. This aligns with the literature, which shows that physicians overwhelmingly desired parallel medical education in both conventional and CAIM topics in order to maintain a broad medical perspective,^20,74^ and yet, were unclear on their responsibilities in discussing CAIM, and prioritized the biomedical model over holistic care _models._21,75,76

#### Distrust and Concern About CAIM Safety and/or Efficacy

Physicians’ concerns about the safety and efficacy of CAIM interventions was identified as a central theme, underpinned by the perceived absence of rigorous research and evidence-based findings. This skepticism often led physicians to prioritize conventional medicine over CAIM and adopt a cautious approach to avoid misleading patients.^21,77^ On the other hand, very few providers are able to elaborate on the mechanism, specifics of the interaction, or the potential dangers of CAM use.^78^ This gap of knowledge among providers on the safety and efficacy of CAM can further drive the stigma of CAM and its divide with conventional medicine.^78^

#### Accepting CAIM as an Important Part of Cancer Care

Similar to our findings, the literature shows that physicians generally believe that CAIM is clinically relevant in cancer care given its potential to impact or interact with conventional treatments.^15,21^ Physicians also agree that incorporating CAIM therapies would have a positive impact on patient satisfaction and autonomy and promotes values of holistic care.^15,70,77^ This is especially important for cancer patients and survivors who are often uncertain about the next steps in their cancer journey and experience significant emotional, spiritual, social, and lifestyle changes.^79^ Holistic care can promote better adherence to treatment plans, and result in better disease outcomes.^80^

#### Communication Dynamics Between Patients and Physicians

A recent review echoed our findings that nondisclosure of CAIM use is common throughout the cancer care spectrum and physicians are often unaware of CAIM use, despite it being highly prevalent among patients.^15^ Patients do not disclose CAIM use due to fears of negative reactions and being advised to stop,^11,15^ while physicians do not initiate discussions due to perceived lack of knowledge on the topic.^21^ Physicians also face structural barriers such as a shortage of time during consultation sessions as well as a lack of knowledge on referral pathways to CAIM providers.^15,76^ It is crucial that physicians initiate discussions about CAIM with patients. Patients put time, money, energy, and hope into CAIM interventions, and physicians should monitor use to ensure patient safety.^25,35^ Physicians’ shared-decision making on CAIM, including maintaining respectful communication and an open-minded attitude,^81^ can also enhance patients’ satisfaction and increase overall compliance and adherence to treatment regimens.^23,24^

### Implications and Future Directions

This study provides insight into the knowledge and experiences of physicians counselling about CAIM in the context of cancer care. The findings emphasize that CAIM is an important part of person-centered and holistic cancer care and there is a need for more rigorous and evidence-based guidelines for physicians counselling about CAIM. Better education and training for physicians is needed to empower them to initiate discussions about CAIM with their patients. It is also important to address administrative and structural barriers to counselling about CAIM such as limited time during clinical appointments and the lack of established referral pathways to CAIM programs. Future research should investigate the knowledge and experience of medical students and trainees to inform educational interventions about CAIM. More research is also needed about the knowledge and experiences of other conventional health providers such as nurses and how they approach CAIM discussions in collaboration with physicians.

#### Strengths and Limitations

A major strength of this review was the comprehensive literature search that was conducted across multiple academic databases, supplemented by hand-searching the reference lists of other relevant review articles. We also adhered to the PRISMA 2020 reporting guidelines, and screening, data extraction, and thematic analysis stages were completed independently and in duplicate. Additionally, all stages were piloted with a small sub-set of articles to ensure consistency and rigour. One limitation is that only English language publications were considered for inclusion and our findings therefore may represent only a subset of cultural understanding and practice. This is especially relevant because CAIM may be practiced more frequently in non-English speaking regions of the world, such as traditional Chinese medicine in China. Further, the included studies originated predominantly from Western countries of Canada, Germany, or Australia. This may limit the generalizability of findings to regions with particular healthcare systems or cultural contexts. Finally, most studies relied on semi-structured interviews, which are susceptible to biases including the potential for the researcher to misinterpret participants’ responses, or inadvertently influence the participant to respond in a particular way. For example, physician participants may respond to interviewers who are physicians differently than those who are not physicians.

## Conclusions

In this systematic review of qualitative studies, we identified 35 eligible studies presenting the knowledge and experiences of physicians counselling about CAIM for cancer care. Four main themes were identified: lack of knowledge and formal training on CAIM; distrust and concern about CAIM safety and/or efficacy; accepting CAIM as an important part of cancer care; and the communication dynamics between patients and physicians. Our findings underscore the need for better education and training to promote CAIM knowledge among physicians and empower them to initiate discussions on CAIM with patients. Our findings also emphasize the need to address administrative and structural barriers for physicians counselling on CAIM in the context of cancer care.

## List of Abbreviations

CAIM: complementary, alternative, and integrative medicine CASP: Critical Appraisal Skills Programme PRISMA: Preferred Reporting Items for Systematic Reviews and Meta-Analyses SPIDER: Sample, Phenomenon of interest, Design, Evaluation, and Research type

## Declarations

### Ethics Approval and Consent to Participate

This study involved a systematic review of peer-reviewed literature only; it did not require ethics approval or consent to participate.

### Consent for Publication

All authors consent to this manuscript’s publication.

### Data Availability

All study materials and data are included in this manuscript or posted on Open Science Framework: https://doi.org/10.17605/OSF.IO/UQ39R

### Author Contributions

Jeremy Y. Ng, MSc, PhD (Conceptualization; Formal Analysis; Investigation; Methodology; Project Administration; Writing – Original Draft; Writing – Review & Editing; Supervision), Aimun Q. Shah, BSc (Conceptualization; Formal Analysis; Investigation; Project Administration; Visualization; Writing – Original Draft; Writing – Review & Editing), Lana Abu Narr, BSc (Formal Analysis; Investigation; Visualization; Writing – Review & Editing), Alyssa Qian (Formal Analysis; Investigation; Visualization; Writing – Review & Editing), Holger Cramer, PhD (Investigation; Methodology; Supervision; Writing – Review & Editing).

## Supplementary Files Legend

Supplementary File 1: Bibliographic Database Search Strategies

Supplementary File 2: General Characteristics of Included Qualitative Studies on Physicians’ Knowledge and Experiences Counseling on Complementary, Alternative, and Integrative Medicine (CAIM) in Cancer Care (N=35)

Supplementary File 3: Outcomes and Findings of Included Qualitative Studies on Physicians’ Knowledge and Experiences Counselling on Complementary, Alternative, and Integrative Medicine (CAIM) in Cancer Care (N=35)

