## Supplementary File 1 for "Physicians’ Knowledge and Experiences of Counseling on Complementary, Alternative, and Integrative Medicine in Cancer Care: A Qualitative Systematic Review"

### **Supplementary File 1:** Bibliographic Database Search Strategies

MEDLINE Search Strategy for Investigating Physicians' Knowledge and Experiences Counselling on Complementary, Alternative, and Integrative Medicine in the Context of Cancer Care, Executed May 25, 2023.

**Database: Ovid MEDLINE(R) and Epub Ahead of Print, In-Process & Other Non-Indexed Citations, Daily and Versions(R) <1946 to May 25, 2023>**
**Search Strategy:**

**1** <CAIM Search String Informed by Operational Definition>.ti. (2472441)
**2** exp Complementary Therapies/ or exp Integrative Medicine/ (245447)
**3** (doctor* or physician* or oncologist*).mp. (763557)
**4** exp Oncologists/ or exp Physicians/ (176102)
**5** (oncology or cancer*).mp. (2323113)
**6** (exp Medical Oncology/ or exp Neoplasms/ (3843820)
**7** (qualitative* or survey* or focus group* or interview* or descriptive* or mixed method*).mp. (1980595)
**8** exp Qualitative Research/ (81547)
**9** 1 or 2 (2625883)
**10** 3 or 4 (796807)
**11** 5 or 6 (4497975)
**12** 7 or 8 (1980745)
**13** 9 and 10 and 11 and 12 (1434)
**14** limit 13 to English language (1368)

**S2.** EMBASE Search Strategy for Investigating Physicians' Knowledge and Experiences Counselling on Complementary, Alternative, and Integrative Medicine in the Context of Cancer Care, Executed May 25, 2023.

**Database: Ovid MEDLINE(R) and Epub Ahead of Print, In-Process & Other Non-Indexed Citations, Daily and Versions(R) <1946 to May 25, 2023>**
**Search Strategy:**

**1** <CAIM Search String Informed by Operational Definition>.ti. (2770748)
**2** exp alternative medicine/ or exp integrative medicine/ (78882)
**3** (doctor* or physician* or oncologist*).mp. (1094203)
**4** exp oncologist/ or exp medical oncologist/ or exp physician/ (970041)
**5** (oncology or cancer*).mp. (4622526)
**6** exp oncology/ or exp neoplasm/ (5556360)
**7** (qualitative* or survey* or focus group* or interview* or descriptive* or mixed method*).mp. (2838659)
**8** exp qualitative research/ or exp qualitative analysis/ or exp semi structured interview/ (232117)
**9** 1 or 2 (2814364)
**10** 3 or 4 (1576912)
**11** 5 or 6 (6374129)
**12** 7 or 8 (2838887)
**13** 9 and 10 and 11 and 12 (2988)
**14** limit 13 to English language (2886)

AMED Search Strategy for Investigating Physicians' Knowledge and Experiences Counselling on Complementary, Alternative, and Integrative Medicine in the Context of Cancer Care, Executed May 25, 2023.

**Database: AMED (Allied and Complementary Medicine) <1985 to May 2023>**
**Search Strategy:**

**1** <CAIM Search String Informed by Operational Definition>.ti. (2472441)
**2** exp Complementary therapies/ or exp Complementary medicine/ or exp Integrative Medicine/ (71149)
**3** (doctor* or physician* or oncologist*).mp. (10644)
**4** exp Physicians/ (1050)
**5** (oncology or cancer*).mp. (10644)
**6** exp Neoplasms/ (17878)
**7** (qualitative* or survey* or focus group* or interview* or descriptive* or mixed method*).mp. (31260)
**8** 1 or 2 (110044)
**9** 3 or 4 (10644)
**10** 5 or 6 (21897)
**11** 7 and 8 and 9 and 10 (96)
**12** limit 11 to English language (90)
