## Supplementary File 2 for "Physicians’ Knowledge and Experiences of Counseling on Complementary, Alternative, and Integrative Medicine in Cancer Care: A Qualitative Systematic Review"

**Supplementary File 2:** General Characteristics of Included Qualitative Studies on Physicians' Knowledge and Experiences Counseling on Complementary, Alternative, and Integrative Medicine (CAIM) in Cancer Care (N=35)

| Title | Author, year of publication | Country | Study objectives | Qualitative methodologies used | Theoretical underpinning of qualitative methodology | Sample characteristics | Themes discussed |
|---|---|---|---|---|---|---|---|
| Are We Meeting Cancer Patient Needs? Complementary and Alternative Medicine Use Among Saudi Cancer Patients: A Qualitative Study of Patients and Healthcare Professionals' Views | Alqahtani et al., 2018 [35] | Saudi Arabia | To examine the perspectives of cancer patients and healthcare professionals' on complementary and alternative medicine in tertiary hospitals that do not provide CAIM. | Semi-structured Interviews | Interpretive Analysis | *N = 30 healthcare professionals, of which n=8 were physicians (also included a patient comparison group), 20% male (n=6), 80% female (n=24), age range 29-47 years, 86.7% had equal to or less than 10 years of experience with cancer care. | 1. Healthcare providers' negative opinions on CAIM use for cancer care<br>2. Healthcare providers' positive opinions on CAIM use for cancer care |
| Patient and Medical Oncologists' Perspectives on Prescribed Lifestyle Intervention—Experiences of Women with Breast Cancer and Providers | Balneaves et al., 2020[36] | Canada | To explore the perspectives and experiences of medical oncologists in referring breast cancer patients to exercise and nutrition programming at a tertiary cancer treatment center. | Semi-structured Interviews | Descriptive Analysis | N= 8 physicians including n=7 medical oncologists and n=1 GP in oncology (also included a patient comparison group), 25% male (n=2), age 30-59, 50% had ≥ 15 years of practice experience. | 1. Perceptions of the prescription of exercise/nutrition programs.<br>2. Benefits of lifestyle intervention. Sub-themes: reframing disease as health, coping with the physical consequences of cancer and beyond, managing distress, gaining social support.<br>3. Perceived challenges of the lifestyle intervention. |

| | | | | | | | |
|---|---|---|---|---|---|---|---|
| "Part of the Conversation": A Qualitative Study of Oncology Healthcare Professionals' Experiences of Integrating Standardized Assessment and Documentation of Complementary Medicine | Balneaves & Watling, 2022[37] | Canada | To explore the experiences and perceptions of oncology healthcare professionals regarding their implementation of CM guideline recommendations, as well as barriers and facilitators they experienced in assessing, documenting, and discussing CM use during clinical practice. | Interviews (one-on-one or in dyads) | Interpretive descriptive analysis | *N= 20 HCPs of which n=4 were medical oncologists, 20% male (n=4), 80% female (n=16), age range 30-50 years, 25% had 1-4 years experience, 15% had 5-10 years experience, and 60% had 10+ years of experience. | 1. Motivations for study participation and withdrawal 2. Past experiences in addressing CM use 3. Experiences of implementing standardized CM assessment and documentation 4. Barriers and facilitators to assessment and documentation of CM use 5. Future intentions related to assessing and documenting CM use 6. Recommendations related to assessment and documentation of CM use. |
| Supportive Care and Osteopathic Medicine in Pediatric Oncology: Perspectives of Current Oncology Clinicians, Caregivers, and Patients | Belsky et al., 2021[38] | USA | To investigate clinician, caregiver, and patient knowledge of osteopathic medicine in the pediatric oncology population and to explore the perception of possible utilization and barriers to OMT. | Semi-structured Interviews (as part of larger mixed method study) | Not reported | *N=20 oncology clinicians of which n=15 were attending physicians, 35% male (n=7), 65% female (n=13), median 7 years of clinical practice (1-24 years). | 1. Patients have uncontrolled chemotherapy side effects. 2. Desire for better supportive care options *Subthemes*: Lack of alternative therapies; desire non-pharmacological interventions 3. OMT is a favorable supportive care option. *Subthemes:* Beneficial intervention - low risk, non-invasive, and non-pharmacologic; Receptive |

| | | | | | | | |
|---|---|---|---|---|---|---|---|
| Communication and Integration: A Qualitative Analysis of Perspectives Among Middle Eastern Oncology Healthcare Professionals on the Integration of Complementary Medicine in Supportive Cancer Care | Ben-Arye et al., 2015[39] | Israel | To explore the perspectives of Middle Eastern oncology healthcare professionals (HCPs) regarding the integration of CTM within conventional supportive cancer care | Questionnaire (mixed methods) | Narrative Analysis | *N=178 oncology HCPs of which n=110 were physicians, 46% male (n=82), 54% female (n=95), mean age 41.5, 75% Muslim, 13% Jewish, 11% Christian, 1% Druze. | 1. HCP related barriers. Subthemes: lack of knowledge of CTM; lack of training in CTM consultation; skeptical attitudes<br>2. HCP-patient communication barriers. Sub-themes: Non-disclosure of CTM use induced by judgmental HCP's attitudes or patient's concerns to reveal non-conventional use; failure to establish realistic expectations of CTM objective; difficulty respecting patient's CTM-related health belief model; unmatched awareness of CTM risks (e.g. herb-chemotherapy interactions); consultations regarding CTM modalities with limited research evidence<br>3. Patient and Caregiver related barriers. Subthemes: Lack of knowledge; unrealistic expectations of CTM effectiveness; unawareness of risks; gaps between EBM and charlatanry<br>4. Barriers outside the hospital. Subthemes: public unawareness of CTM risks and overstated effectiveness claims; lack of CTM legislation |

| | | | | | | | |
|---|---|---|---|---|---|---|---|
| Physicians' Attitudes Toward Patients' Use of Alternative Cancer Therapies | Bourgeault, 1996[40] | Canada | To determine physicians' attitudes and reactions to their patients' use of alternative cancer therapies, factors that affect these reactions, and physicians' views of how the use of such therapies affects the physician-patient relationship. | Semi-structured interviews | Grounded theory | N=30 physicians of which n=18 were oncologists and n=12 general practitioners, 76.67% (n=23) male, n=10 < 40 years of age, n=16 between 40-60 years, and n=4 >60 years. | 1. Familiarity with alternative cancer therapies.<br>2. Attitudes toward efficacy of alternative cancer therapies.<br>3. Reactions to patients' use of alternative cancer therapies.<br>4. Effect on physician-patient relationships. |
| Oncology Clinicians' Accounts of Discussing Complementary and Alternative Medicine With Their Patients | Broom & Adams, 2009[41] | Australia | To explore oncologists' self-reported approaches to discussions about CAIM with their patients, team dynamics, and professional education; and to investigate oncologists' knowledge of CAIM. | Interviews | Interpretive Analysis | *N=25 oncology HCPs of which n=13 were oncologists. | 1. Oncologists on discussing CAIM with patients<br>2. Protecting the patient or protecting the doctor Representing risk to the patient<br>3. Why do our patients use CAIM? Psychosocial support and terminality<br>4. Communication, lack of information and physician culture<br>5. Education and CAIM |

| | | | | | | | |
|---|---|---|---|---|---|---|---|
| Traditional Medicines, Collective Negotiation, and Representations of Risk in Indian Cancer Care | Broom & Doron, 2012[42] | India | To examine Indian oncologists' experiences of communicating with patients on TCAM. | Semi-structured Interviews | Interpretive analysis | *N=22 clinicians of which n=16 were physicians, 55% male (n=12), 45% female (n=10). | 1. Cancer, Contagions, and biomedical literacy<br>2. Individualism, Collectivism, and Relationism<br>3. TCAM, Nondisclosure, and Dissuasion<br>4. The Rhetoric and Realities of Risk and Impurity |
| Bridging the Gap Between Attitudes and Action: A Qualitative Exploration of Clinician and Exercise Professional's Perceptions to Increase Opportunities for Exercise Counseling and Referral in Cancer Care | Caperchione et al., 2022[43] | Australia | To understand barriers and facilitators impacting the implementation of exercise communication and referral, and to explore integrated clinical approaches to exercise communication and referral in cancer care. | Focus groups | Descriptive (naturalistic approach) | *N=39 cancer care clinicians (medical/radiation oncologists, oncology nurses, psychologists, and social workers) across 5 focus groups (also included exercise professionals in 2 focus groups). | 1. Factors impacting knowledge to action gap. *Sub-themes*: Lack of knowledge and education; funding structure; and current referral pathway.<br>2. Recommendations for a consistent and efficient way forward. *Sub-themes*: Early and ongoing communication about exercise; tailored communication and resources for oncologists and patients; and exercise practitioner network and referral path. |
| Changing Physicians' Attitudes Toward Self-help Groups: An Educational Intervention | Carroll et al., 2000[44] | Canada | To assess family physicians' attitudes towards self-help groups, and to examine whether these attitudes could be changed through an educational intervention. | Questionnaire (mixed-method) | Not reported | N= 584 family physicians (35% or n=204 of which provided open-ended comments), 42.7% female and 57.3% male, mean years 11.95 (0-40) in active practice. | 1. Awareness of and practices related to self-help groups.<br>2. Attitudes (and attitude shifts) towards self-help groups. |

| | | | | | | | |
|---|---|---|---|---|---|---|---|
| Oncologists' Experiences of Discussing Complementary and Alternative Treatment Options With Their Cancer Patients. A Qualitative Analysis | Corina et al., 2016[45] | Germany | To investigate real-world discussions of CAM treatments between physicians and patients by learning about the values, norms, and defining features that impact this communication. | Semi-standardised interviews | Interpretive analysis | N=17 oncologists (n=11 working in inpatient settings and n=6 in outpatient settings), 59% males (n=10), 41% females (n=7), ages (33-58 years), qualified to practice (6-33 years). | 1. CAIM as a treatment option from 'another world'<br>2. 'Advice as a service'<br>3. 'Role of knowledge and evidence' |
| Complementary Medicine for Cancer Patients in General Practice: Qualitative Interviews with German General Practitioners | Dahlhaus et al., 2015[46] | Germany | To investigate general practitioners' reactions when cancer patients display interest in complementary medicine and when asked about their knowledge in the field. | Semi-structured Interviews | Inductive and deductive methods, Mayring's qualitative content analysis | N= 10 GPs including n=7 family physicians, n=2 internal medicine physicians, n=1 family and internal medicine physicians, 40% male (n=4), 60% female (n=6), experience ranged from 5-41 years, 60% had 5-10 years of professional experience, 30% had 11-40 years of professional experience, 10% had 40+ years of professional experience. | 1. GPs Feel Responsible for Advising Patients on the Usefulness of CAIM<br>2. GPs Express the Need for More Education and Information on CAIM<br>3. What Advice Should You Provide When You Lack Knowledge? |
| A Qualitative Study of Patient and Healthcare Provider Perspectives on Building Multiphasic Exercise Prehabilitation | Daun et al., 2022[47] | Canada | To receive feedback from HCPs and HNC surgical patients on incorporating exercise into standard HNC surgical care. | Semi-structured interviews | Interpretive description, constructivist | *N=10 healthcare providers of which n=4 were physicians/surgeons (also included a patient comparison group), 40% male (n=4), 60% female (n=6). | 1. Assessments are acceptable and necessary. Subthemes: challenges; strengths<br>2. Value of exercise and its importance in clinical care. Subthemes: perceptions of exercise for physical and psychosocial outcomes; |

| | | | | | | | |
|---|---|---|---|---|---|---|---|
| into the Surgical Care Pathway for Head and Neck Cancer | | | | | | | perceptions of exercise in-hospital<br>3. Components of an ideal multiphasic exercise pre-habilitation program. Subthemes: need for individualization; frequency, intensity, time and type (FITT) principles<br>4. Key factors will support implementation. Subthemes: education; HCP role; need for a culture shift in cancer care |
| The Challenge of Timing: A Qualitative Study on Clinician and Patient Perspectives About Implementing Exercise-Based Rehabilitation in an Acute Cancer Treatment Setting | Dennet et al., 2020[48] | Australia | To assess barriers and facilitators of implementing an exercise rehabilitation program for cancer survivors in an acute setting. | Semi-structured interviews (focus group and individual) | Phenomenological | *N=25 clinicians of which n=4 were physicians including 3 oncologists and 1 hematologist (also included a patient group), 12% male (n=3), 88% female (n= 22), mean 11.4 years of oncology experience. | 1. Finding the 'right time' for rehabilitation. Subthemes: attitude, knowledge, convenience, resources |
| Physician Perspectives on Unconventional Cancer Therapies / A Comparison of Physician and Patient Perspectives on Unconventional Cancer Therapies | Gray et al., 1997; Gray et al., 1998[49,50] | Canada | To assess physician and cancer survivor perspectives and reactions to unconventional cancer therapies and to focus more broadly on challenges related | Interviews with Open-ended questions | Not reported | *N = 19 physicians including n=9 general practitioners, n=9 oncologists, n=1 surgeon (also included a patient group), 36.8% male (n=7), 63.2% female (n=12). | 1. The need for information: Attitudes towards information provision and physician roles<br>2. What is unconventional? Who uses unconventional therapies and reasons for their popularity<br>3. How do physicians respond to patient interest in unconventional therapies? |

| | | | | | | | |
|---|---|---|---|---|---|---|---|
| | | | to unconventional therapies. | | | | 4. What is the ideal interface between practitioners of conventional and unconventional therapies?<br>5. Why are there communication problems between patients and physicians about unconventional therapies?<br>6. What criteria are legitimate for making decisions about unconventional therapies?<br>7. Physicians' attitudes towards and feelings about unconventional therapies |
| What Hinders Healthcare Professionals in Promoting Physical Activity Towards Cancer Patients? The Influencing Role of Healthcare Professionals' Concerns, Perceived Patient Characteristics and Perceived Structural Factors? | Haussmann et al., 2018[51] | Germany | To provide a comprehensive description of factors that influence HCPs' promotion of PA to patients with cancer, and to identify the reasons, motives, and mechanisms behind these factors. | Semi-structured interviews | Not reported | *N= 30 HCPs of which n=20 are physicians, including n=10 GPs and n=10 specialized physicians, 37% male (n=11), 63% female (n=19), mean age 45 years, mean 34.8 cancer patients treated per month. | 1. HCP's concerns. Subthemes: physical overexertion of patients, psychological stress for patients.<br>2. Perceived patient characteristics. Subthemes: Patient's physical condition, patient's assumed interest in PA, patient's former PA level.<br>3. Perceived structural factors. Subthemes: HCPs' workload, in-house structures, timing and coordination, information available, and the availability of the exercise program. |

| | | | | | | | |
|---|---|---|---|---|---|---|---|
| On Caring and Sharing–Addressing Psychological, Biographical, and Spiritual Aspects in Integrative Cancer Care: A Qualitative Interview Study on Physicians' Perspectives / The Subjective Dimension of Integrative Cancer Care: A Qualitative Study Exploring the Perspectives, Themes, and Observations of Experienced Doctors from the Area of Anthroposophic Medicine | Kienle et al 2018a; Kienle et al 2018b[52,53] | Germany | To explore what role psychological, biographical, and spiritual factors play for experienced doctors working in integrative cancer care; to explore perceptions, themes, goals, procedures, and observations of experienced AM doctors with regard to the subjective dimensions of ICC. | Semi-structured interviews | Not reported | N=35 physicians including n=17 in internal medicine, pulmonology, or gastroenterology, n=12 GPs, n=8 oncologists and hematologists, n= 3 pediatricians, n=1 gynecologist, n=1 neurologist, and n=1 researcher, 85.7% male (n=30), 14.3% female (n=5), mean age 55 years (40-84), mean 26 years of physician work experience, 21 working in outpatient clinic or hospital and 14 resident doctors. | 1. Doctors and treatment setting<br>2. Concepts, themes and treatment goals<br>3. Strengthening, vitality, warmth and recovery<br>4. Regeneration, relief from suffering<br>5. Living a normal life and finding one's own way. Subthemes: Autonomy, stabilization, and coping; feeling emotionally secure<br>6. Triangulation with published studies on patients' perspective<br>7. Spirituality. Subthemes: Characteristics of spirituality; talking about spiritual issues; spiritual attitude of doctors; acceptance and emotional freedom from cancer |
| German Physicians' Perceptions and Views on Complementary Medicine in Pediatric Oncology: A Qualitative Study | Klatt et al., 2023[54] | Germany | To explore German POs' understanding of CAIM, related attitudes, and challenges and strategies related to CAIM discussions. | Semi-structured interviews | Thematic analysis, interpretive | N=14 POs, 50% male (n=7), 50% female (n=7), mean age 30.8 years (33-54), mean years of experience 13.8 years (0.5-26), 10 worked in university hospitals and 4 worked in non-university hospitals; 7.1% was qualified in natural therapies, 7.1% was qualified in | 1. POs' understanding of CAIM<br>2. Perceived benefits and harms of CAM<br>3. Counseling on CAIM: rationale, barriers, and strategies |

| | | | | | | | |
|---|---|---|---|---|---|---|---|
| | | | | | | rehabilitation medicine, 7.1% was qualified in pain therapy, 28.6% was qualified in palliative medicine, and 14.9% was qualified in other. | |
| Lifestyle Advice to Cancer Survivors: A Qualitative Study on the Perspectives of Health Professionals | Koutoukidis et al., 2017[69] | United Kingdom | To explore HPs' views on counseling on lifestyle advice (e.g., healthy eating, PA, smoking, alcohol) for cancer survivors. | Semi-structured interviews | Not reported | *N= 21 HPs of which n=11 are physicians, including n=4 surgeons, 24% male (n=5), 76% female (n=16), 5% were 26-35 years old, 38% were 36-45 years old, 48% were 46-55 years old, 10% were 56-65 years old, 43% specialized in breast cancer, 38% specialized in colorectal cancer, 19% specialized in prostate cancer. | 1. HPs' perceptions of survivor-centered barriers to provision of lifestyle advice. Subthemes: Perceptions of survivors' current health behaviours and ability to perform health behaviours, potential loss of connection with the patient, socioeconomic barriers<br>2. HP-centered barriers to provision of lifestyle advice. Subthemes: Knowledge and attitudes towards evidence and guidelines, self-identification as the right person to provide lifestyle advice, practical barriers to lifestyle advice provision.<br>3. Optimal delivery of lifestyle advice. Subthemes: Tailored advice delivered during cancer care, small and achievable changes framed as part of treatment<br>regimen, cost-effectiveness of scalability |

| | | | | | | | |
|---|---|---|---|---|---|---|---|
| Academic Doctors' Views of Complementary and Alternative Medicine (CAM) and Its Role Within the NHS: An Exploratory Qualitative Study | Maha & Shaw, 2007[55] | United Kingdom | To investigate the views and accompanying rationales of academic doctors on CAIM and its role within the NHS. | Semi-structured interviews | Pragmatic, subtle realist perspective | *N=9 healthcare providers of which n=8 are GPs. | 1. The role of doctors' professional experiences in shaping their views of CAIM.<br>2. Doctor-patient communication about CAM and patient disclosure.<br>3. Training and education in CAIM: is there a need?<br>4. Hierarchy of acceptability of CAM and the nature of evidence<br>5. The role of CAIM within the NHS. |
| Yoga in Adult Cancer: A Pilot Survey of Attitudes and Beliefs Among Oncologists | McCall & Heneghan, 2015[56] | Canada | To (i) test the acceptability of a self-reported online survey for oncologists; (ii) generate preliminary evidence on oncologists' knowledge, attitudes, beliefs, and current referral practices regarding yoga for adult cancer patients; and (iii) to list the perceived benefits and barriers to yoga from a clinical perspective. | Self-report questionnaire (mixed methods) | Not reported | N=29 oncologists including 17 medical oncologists and 12 radiation oncologists, 52% male (n=15), 48% female (n-14), mean age 41.9 years (28-56), 41% had less than 5 years of experience, 45% had 5-25 years of experience, 45% had 5-25 years of experience, 14% had more than 25 years of experience, 62% White, 17% Chinese, 17% Indian, 3% Hispanic. | 1. Attitudes Toward Yoga<br>2. Perceptions of Yoga Evidence<br>3. Knowledge and Beliefs About Yoga<br>4. Patterns of Yoga Use in Conventional Treatment<br>5. Perceived Barriers to Yoga Adherence |

| | | | | | | | |
|---|---|---|---|---|---|---|---|
| How is Complementary Medicine Discussed in Oncology? Observing Real-Life Communication Between Clinicians and Patients with Advanced Cancer | Mentink et al, 2022[57] | Netherlands | To examine the structure of communication about CM between cancer patients and clinicians during oncology consultations. | Secondary analysis of audio/video recordings of consultations | Not reported | *N=29 healthcare professionals, including 13 medical oncologists and 9 radiation oncologist (also included a patient group), 35% male (n=10), 65% female (n=19). | 1. How is CM introduced<br>2. Clinicians' response to introduction of CM.<br>3. Mentioned aspects that are related to CM. Subthemes: safety, effectiveness, and costs of CM, alternative options for CM use, contact with CM provider, information source and alternative treatment, and patient choice.<br>4. Clinician attitude towards CM |
| Oncologists' and Naturopaths' Nutrition Beliefs and Practices | Novak & Chapman, 2001[58] | Canada | To explore beliefs of physicians and CAIM practitioners, their use of scientific and other types of evidence, and their counseling practices, specifically regarding the role of diet in breast cancer prevention and treatment. | Semi-structured interviews | Blurred genres, taking a pragmatic approach | *N=10 oncologists of which 6 were radiation oncologists and 4 were medical oncologists (also included naturopath group), 80% male (n=8), 20% female (n=2), 80% White, 20% Asian, mean 15.4 years of clinical experience. | 1. Beliefs regarding diet and breast cancer etiology<br>2. Evidence regarding diet and breast cancer<br>3. Dietary counseling practices and breast cancer |
| Complementary Therapy Use by Cancer Patients: Physicians' Perceptions, Attitudes, and Ideas | O'Beirne et al., 2004[59] | Canada | To explore family physicians' perceptions of their cancer patients' use of complementary therapy. | Focus groups | Not reported | N=28 family physicians, 75% male (n=21), 25% female (n=7), ages 35-50 years, 43% from rural settings, 57% from urban settings. | 1. Definition of complementary therapies<br>2. Importance of holistic health<br>3. Role of evidence<br>4. Attitudes toward complementary therapies |

| | | | | | | | |
|---|---|---|---|---|---|---|---|
| | | | | | | | 5. Perceptions of use of complementary therapies<br>6. Patient-physician communication<br>7. Family physicians' role with complementary therapies<br>8. Concerns about complementary therapies |
| Perspectives of Pediatric Oncologists and Palliative Care Physicians on the Therapeutic Use of Cannabis in Children with Cancer | Oberoi et al., 2022[60] | Canada | To understand the perspectives and practices of pediatric oncologists and palliative care physicians on the use of medical cannabis for pediatric cancer patients and to determine the necessity for cannabis-related research. | Survey (mixed methods) | Not reported | *N =119 HCPs, of which n=94 were physicians and 25 were fellows, 50.4% from central Canada, 40.4% from western Canada, 9.2% from eastern Canada. | 1. Lack of high quality research<br>2. Patient's and parents' beliefs<br>3. Need for high quality studies<br>4. Misinformation from the online content<br>5. Patient engagement and informed decision making<br>6. Policies around use of cannabis<br>7. Possible harms<br>8. Financial burden |
| Convergent Priorities and Tensions: a Qualitative Study of the Integration of Complementary and Alternative Therapies with Conventional Cancer Treatment | River et al., 2018[61] | Germany | To examine HCPs' and patient dynamics in an integrated cancer service where CAIM is provided alongside standard cancer treatments, and to understand drivers or barriers to further | Semi-structured interviews | Interpretive-description | *N=7 HCPs of which n=3 were oncologists, aged >18 years, all English speaking. | 1. Prioritizing person-centered care<br>2. What constitutes evidence?<br>3. Who should bear the costs? |

| | | | | | | | |
|---|---|---|---|---|---|---|---|
| | | | integration of CAIM with standard cancer treatment. | | | | |
| Mind the Gap! Lay and Medical Perceptions of Risks Associated with the Use of Alternative Treatment and Conventional Medicine | Salamonsen, 2015[62] | Norway | To explore lay and medical risk perceptions associated with CAIM and conventional medicine. | Interviews | Exploratory, content analysis | N=12 physicians including n=4 oncologists, n=3 neurologists, n=5 general practitioners (also included a patient group), 50% (n=6) male, age range 41-65, over 10 years of practice experience, 2 GPs were also experienced CAIM practitioners. | Not applicable |
| 'They Don't Ask Me So I Don't Tell Them': Patient-Clinician Communication About Traditional, Complementary, and Alternative Medicine | Shelley et al., 2009[63] | USA | To compare perspectives of patients and primary care clinicians on communication about TM/CAIM, and identify strategies to enhance patient-clinician communication. | Semi-structured interviews (focus groups and individual) | Grounded theory: editing analytic approach, immersion/crystallization | *N=19 clinicians of which n=16 are physicians, including n=13 family physicians, n=1 pediatrician and n=2 internists, 62.5% male (n=10), 37.5% female (n=6). | 1. Acceptance/non-judgment<br>2. Initiation of communication<br>3. Safety/efficacy concerns |

| | | | | | | | |
|---|---|---|---|---|---|---|---|
| Attitudes Toward Complementary and Alternative Medicine Amongst Oncology Professionals in Brazil | Siegel et al., 2016[64] | Brazil | To provide insights into HCPs' views on the rise, validity, and role of CAIM in cancer care. | Interviews | Not reported | N=32 HCPs of which n=11 are physicians, 28% male (n=9), 72% female (n=23), ages 23-64 years. | 1. Physicians: from encouraging integration to opposition<br>2. Physicians: benevolence but questioning of the viability of integration<br>3. Physicians: dis-interest and/or underlying negativity |
| Considerations in Developing and Delivering a Non-Pharmacological Intervention for Symptom Management in Lung Cancer: The Views of Healthcare Professionals | Wagland et al., 2012[65] | United Kingdom | To explore the views of HCPs involved with cancer care on the most appropriate ways of developing and delivering a non-pharmacological intervention for symptom-management. | Focus groups and telephone interviews | Not reported | N=30 HCPs of which n=4 were oncologists. | 1. Current standard care<br>2.Positive attitudes towards NPIs amongst HCPs<br>3. Non-pharmacological interventions should be personalized and delivered by HCPs<br>4. Factors influencing the effectiveness of non-pharmacological interventions<br>5. Optimum time of intervention delivery<br>6. Delivering non-pharmacological interventions within the community<br>7. Group or individually based teaching of non-pharmacological interventions<br>8. Involving caregivers within the teaching of non-pharmacological interventions |
| 'Probably Better Than Any Medication We Can Give You' | Waterland et al., 2020[66] | Australia | To report GPs' experiences of providing nutrition and exercise advice to cancer patients, and | Semi-structured interviews | Qualitative description, influenced by grounded-theory | N=23 GPs, 52.2% male (n=12), 47.8% female (n=11), 52.5% practiced in metropolitan regions, 47.8% practiced in rural regions, 13% had less | 1. The importance of exercise and nutrition recommendations for cancer patients<br>2. The influence of the patient agenda |

| | | | | | | | |
|---|---|---|---|---|---|---|---|
| | | | identify perceived barriers and facilitators of providing said exercise and nutrition advice. | | | than 5 years of experience, 8.7% had 5-10 years of experience, 78.3% had more than 10 years of experience. | 3. The influence of additional training or personal interests<br>4. Limitations of the primary care setting |
| Communication About Complementary and Alternative Medicine When Patients Decline Conventional Cancer Treatment: Patients' and Physicians' Experiences | Wode et al., 2023[67] | Sweden | To explore patients' and physicians' experiences when patients refuse conventional cancer treatment and turn to using CAIM. | Semi-structured interviews | Interpretive description, framework analysis | *N=10 physicians including n=9 oncologists and n=1 palliative care physician (also included a patient group), 60% male (n=6), 40% female (n=4), mean age 47 years (36-66), mean years of experience 18 years (7-49). | 1. Divergent Perspectives on Treatment Choices. Subthemes: Treatment and Care Addressing Illness and Health, Consequences of Treatment Choices and Costs, Experiences of CAIM and Conventional Treatment, Diverging Views on Treatment Choices<br>2. Roles and Power Struggles in the Patient-Physician Relationship. Subthemes: Roles in Treatment Decisions, Power Struggles and Pushes Toward Extreme Positions<br>3. Paving the Way for Improved Communication in Difficult Treatment Decisions. Subthemes: Transparency and Interest, Respect and Support |
| Medical cannabis: An Oxymoron? Physicians' Perceptions of Medical Cannabis | Zolotov et al., 2018[68] | Israel | To gain a deeper understanding of physicians' views on medical cannabis and its possible integration into their clinic, and to | Semi-structured interviews | Socio-narratology | N=24 physicians including n=9 family physicians, n=6 pain specialist physicians, and n=9 oncologists, 70.8% male (n=17), 29.2% female (n=7), average seniority of 19 | 1. Cannabis as a non-medicine<br>2. Cannabis as a medicine |

| | | | | | | |
|---|---|---|---|---|---|---|
| | | | identify potential underlying factors that influence these perceptions. | | | years (3-33), 91.7% with experience in recommending medical cannabis. |

**Abbreviations**: AM: Anthroposophic Medicine; AT: Alternative Therapy; CAIM: Complementary, Alternative, and Integrative Medicine; CM: Complementary Medicine; CONV: Conventional Medicine; CTM: Complementary and Traditional Medicine; GP: General Practitioner; HCP: Healthcare Professional; HNC: Head and Neck Cancer; HP: Health Professional; ICC: Integrative Cancer Care; MT: Mistletoe Therapy; NPI: Non-Pharmacological Intervention; OMT: Osteopathic Manipulative Treatment; PA: Physical Activity; PCC: Patient Centered Care; PCP: Primary Care Provider; PO: Pediatric Oncologist; PRO: Patient Reported Outcome; QSR: Qualitative Systematic Review; SCO: Supportive Care in Oncology; SEGT: Supportive Expressive Group Therapy; TCAM: Traditional, Complementary, and Alternative Medicine; USA: United States of America

* Studies that include multiple health professionals. Qualitative findings have been limited to physicians only.
