## Supplementary File 3 for "Physicians’ Knowledge and Experiences of Counseling on Complementary, Alternative, and Integrative Medicine in Cancer Care: A Qualitative Systematic Review"

**Supplementary File 3:** Outcomes and Findings of Included Qualitative Studies on Physicians' Knowledge and Experiences Counselling on Complementary, Alternative, and Integrative Medicine (CAIM) in Cancer Care (N=35)

| Title | Author, Year of Publication | Outcomes | Main Findings | Challenges Encountered by Study Population | Limitations | Conclusion |
|---|---|---|---|---|---|---|
| Are We Meeting Cancer Patient Needs? Complementary and Alternative Medicine Use Among Saudi Cancer Patients: A Qualitative Study of Patients and Healthcare Professionals' Views | Alqahtani et al., 2018[35] | Physicians' personal experiences of dealing with CAIM and cancer; physicians' knowledge of CAIM use for cancer; physician communication about CAIM with patients. | 1. Physicians feel restricted by national policy frames which narrow their freedom to speak on CAIM.<br>2. Physicians are concerned with officialising the utilization of CAIM for cancer patients due to the link between CAIM and religious affiliations in the country.<br>3. Physicians are less willing to include CAIM among available healthcare procedures for cancer patients and are less likely to refer patients to CAIM specialists.<br>4. Physicians believe that the medical model should be the priority and standard in providing care; it would be inappropriate to develop medical healthcare policies relating to CAIM. | 1. CAIM policy would conflict with conventional medicine. | 1. Small sample size.<br>2. Population restricted to 3 oncology centres resulting in lower representativeness of physicians in the healthcare provider sample.<br>3. Several types of CAIM are discussed, but there is an absence of an official definition of CAIM in Saudi Arabia.<br>4. Many physicians were unable to be contacted or rejected the request to participate. | In Saudi Arabia, it is insufficient to understand cancer from a solely biomedical perspective. It must also be understood in the context of cultural beliefs about disease etiology and the needs for culturally-appropriate CAIM interventions. Healthcare professionals in Saudi Arabia should be equipped to discuss perceived benefits of CAIM with patients to ensure they are providing patient-centred care. |
| Patient and Medical Oncologists' Perspectives on | Balneaves et al., 2020[36] | Approach to introducing an exercise intervention as part of breast | 1. Prescribing CAIM (i.e. Lifestyle intervention) as opposed to simply suggesting was powerful and taken more | 1. Difficulty in contextualizing the lifestyle intervention to the unique needs of | 1. Small sample size.<br>2. Lack of findings' transferability to other lifestyle interventions | Physicians have shown support for integrating exercise and nutrition |

| | | | | | | |
|---|---|---|---|---|---|---|
| Prescribed Lifestyle Intervention—Experiences of Women with Breast Cancer and Providers | | cancer treatment; perceptions and facilitators; perceived challenges experienced by patients. | seriously by patients and families.<br>2. Physicians suggest that nurses should be front and centre in providing lifestyle recommendations.<br>3. Physicians recognize a need to be more knowledge about nutrition and exercise, and to consistently address lifestyle as a core component of cancer care.<br>4. The lifestyle intervention allowed physicians to reframe disease- or treatment-focused conversations into encouraging ones that promoting health and well-being.<br>5. Physicians had knowledge of compelling evidence that lifestyle interventions are an effective adjunct treatment for cancer to decrease treatment side effects and for weight management.<br>6. Physicians concerned that prescribing the intervention may cause guilt for patients who do not follow the recommendation. Older individuals with pre-existing physical disabilities may also find it difficult to engage. | individuals (e.g., age, fitness goals, work, and family commitments), and lack of customization of the program to patients' SES which may have impacted their engagement. | due to the study's urban setting, using exercise and nutrition programming as a "prescription", and focus on breast cancer.<br>3. Lack of diversity within sample, which mainly included White or Asian, well-educated women with high-SES, reducing the generalizability of results. | programming into standard care, as evident through their experiences with positive patient outcomes and feedback from incorporating such programming into breast cancer patient care. |
| “Part of the Conversation”: A Qualitative | Balneaves & Watling, 2022[37] | Physicians' past and current experiences of standardizing the | 1. Physicians believed that understanding patients' use of CM was important to provide | 1. Lack of education and training on CM therapy (leading to | 1. Potential for selection bias (individuals who agreed to participate re | Considering that many cancer patients use CM |

| | | | | | | |
|---|---|---|---|---|---|---|
| Study of Oncology Healthcare Professionals' Experiences of Integrating Standardized Assessment and Documentation of Complementary Medicine | | assessment and documentation of CM use; barriers and facilitators perceived by physicians in addressing CM use with cancer patients. | holistic, person-centred care, and not understanding patient views could "sabotage" the patient-provider relationship.<br>2. Several physicians were concerned that assessing CM use could be perceived by patients as an endorsement.<br>3. CM assessment occurred sporadically if at all, and not in a systematic or comprehensive manner. It was addressed typically only at the initial consult or when patients brought forward questions.<br>4. Limited assessment of CM use was due to lack of time, insufficient knowledge of CM therapies and their efficacy or side effects, and discomfort in addressing questions about CM.<br>5. Physicians found the process of implementing CM assessment forms generally easy although time constraints were a barrier, and some found the form not user-friendly due to specificity of CM remedy names. There was also concern with lack of follow-up on CM information collected from patients.<br>6. There is a need for evidence-based resources that HCPs and patients can access to learn | discomfort in addressing patient questions).<br>2. Issues in the healthcare system (i.e., lack of standardized assessment procedures, CM therapies not included in the EHR, insufficient time).<br>3. Individual factors, such as attitudes toward CM, perceived lack of knowledge about CM, and fear of legitimizing CM use were challenges in this population.<br>4. Lack of institutional policies, institutional support, and resources for CM. | unique and have polarizing attitudes or experiences toward a subject matter).<br>2. Lack of representativeness in the sample reducing generalizability to all HCPs as recruitment is from a single cancer center.<br>3. May not have reached data saturation due to limited sample size and the complexity of CM use. | interventions, oncology HCPs believe that addressing CM use is an essential and feasible component of high-quality, person-centered cancer care. Despite this, there are institutional and professional challenges that must be addressed before assessment, documentation, and discussion of CM during physician-patient consultations can be improved. |

| | | | | | | |
|---|---|---|---|---|---|---|
| | | | about CMs and encourage dialogue. | | | |
| Supportive Care and Osteopathic Medicine in Pediatric Oncology: Perspectives of Current Oncology Clinicians, Caregivers, and Patients | Belsky et al., 2021[38] | Knowledge of osteopathic manipulative treatments (OMT); satisfaction level with current supportive care and options; level of interest in having OMT available during chemotherapy, once educated; how they felt OMT could be best introduced to their patients. | 1. Physicians expressed frustration with currently available treatments including insufficiency of multiple pharmacologic interventions and a lack of alternative available options and reported similar frustration in families of patients.<br>2. Physicians felt inadequate when approached by patients and caregivers who desired non-pharmacologic interventions for conventional treatment side effects due to lack of knowledge.<br>3. Once educated on OMT, physicians did not hesitate in referring patients for OMT<br>4. Need for additional research on osteopathic medicine in pediatric oncology patients to demonstrate efficacy and feasibility in clinic setting. | 1. Lack of knowledge on CAIM.<br>2. Lack of OMT practiced by osteopathic physicians at large pediatric institutions compared to rural adult hospitals<br>3. Lack of osteopathic research within the field, which lays the groundwork for future feasibility and efficacy supportive care clinical trials for OMT in pediatric oncology patients | 1. Small sample size; single institution likely reducing the sample representativeness of the wider pediatric oncology population.<br>2. Potential implicit bias due to the lead investigator being an osteopathic physician.<br>3. Potential for observation bias, however, it was mitigated by having two researchers that independently coded the data. | The need for better supportive care options was identified for pediatric oncology patients. Physicians who were educated on OMT portrayed a desire to make OMT available as an adjunct treatment. This study identified a gap in the research surrounding the feasibility, safety, and efficacy of OMT as a non-pharmacologic supportive care therapy for pediatric cancer patients. Scientifically rigorous clinical trials addressing this question should be conducted in the future. |
| Communication and Integration: A Qualitative Analysis of Perspectives Among Middle Eastern Oncology | Ben-Arye et al., 2015[39] | Perspectives of oncology HCPs on integrating CTM in supportive cancer care; knowledge of CTM amongst physicians. | 1. HCPs' lack of knowledge and training on CTM, in addition to a lack of rigorous research on it, leads to HCP skepticism, creating a barrier to CTM integration.<br>2. From experience in | 1. Low rate of disclosure on CTM use by patients.<br>2. Cross-cultural context in the Middle East, where traditional and high-tech modern | 1. Wording of open-ended questions may have led to a selection bias favoring those who support integration.<br>2. While the study may reflect the cultural | Middle Eastern oncology HCPs welcomed CTM integration within supportive cancer care. However, they suggested the need |

| | | | | | | |
|---|---|---|---|---|---|---|
| Healthcare Professionals on the Integration of Complementary Medicine in Supportive Cancer Care | | | counselling patients, HCPs recognized a need for providing educational resources on CTM to patients, family members, and caregivers.<br>3. HCPs identified that establishing realistic goals for CTM treatment (ex., improving QoL as opposed to curing cancer) is essential during patient/caregiver communication.<br>4. To increase the likelihood of CTM use disclosure, HCPs must understand patients' expectations from CTM and maintain non-judgemental attitudes even if they disagree with patients' beliefs.<br>5. HCPs recognize the need to counsel patients on CTM during early treatment stages. However, they fear that a lack of public awareness and legislation of CTM-related risks would divert patients to charlatans providing fake remedies. | health models interact, poses a challenge with integrating CTM.<br>3. Lack of training and education on CTM, which resulted in a lack of knowledge and skepticism.<br>4. CTM lacked proper regulation, making physicians suspicious of "promising" treatments. 5. Not every response provided sufficient information for thematic analysis. | uniqueness of the Middle East, the work environment of respondents varied greatly, which may affect their responses.<br>3. The small size of the sample, the use of snowball sampling methodology and the lack of homogeneity of the sample precluded further analysis of the data.<br>4. Use of a broad definition of CTM may have affected the response rate, in which the definition may have been seen as inaccurate by European or American readers. | for CTM education and training. A more comprehensive understanding of CTM would provide the knowledge and skills to promote a non-judgmental, evidence-based approach, fostering better patient communication. |
| Physicians' Attitudes Toward Patients' Use of Alternative Cancer Therapies | Bourgeault, 1996[40] | General thoughts and experiences: Familiarity with alternative cancer therapies; experiences with patients' use of alternative cancer | 1. Physicians were relatively unfamiliar with ATs. Sources of information were either their patients, or non-medical sources.<br>2. Use of ATs was considered harmful if it prevented or delayed or interfered with | 1. Lack of familiarity/knowledge of ATs.<br>2. Tension and conflict between patient and provider can occur when the patient is in denial about having | 1. Representativeness of sample. | Physicians lack information on ATs for cancer and believe that most of these therapies have not been scientifically proven. Physicians' attitudes |

| | | | | | | |
|---|---|---|---|---|---|---|
| | | therapies, attitudes and reactions to patients' use of therapies, and the effect of usage on their relationship with patients.<br><br>Additional outcomes after initial analysis: attitudes and behaviours regarding specific circumstances arising from the type of treatment used; prognosis with standard treatment; and the inclusivity of patients' use of alternative therapies. | conventional treatment.<br>3. Physicians believed more evidence is needed to prove efficacy of ATs for it to be adopted as part of standard treatment.<br>4. In situations where no standard treatment is available, physicians are less likely to be so adamantly opposed to the use of ATs.<br>5. If an AT a patient wanted was not harmful, then physicians' behaviour and management of the disease would not change.<br>6. Physicians would neither "forbid" use of the AT, nor "refuse to provide further care" but would stress benefits and encourage standard treatments.<br>7. Some had referred or would refer patients to practitioners of ATs (mainly psychological approaches such as imagery) to deal with side effects of standard therapy.<br>8. Patients' use of ATs can either cause strain, no effect, or positive effects on the physician-patient relationship. | cancer or rejected recommendations of standard medical treatment.<br>3. Physicians often do not know whether a patient is using an AT. | | and reactions to their use by patients are influenced to a greater degree by the efficacy or inefficacy of standard treatment and the invasiveness of the alternative therapy than by the efficacy of the alternative therapy used. |
| Oncology Clinicians' Accounts of Discussing Complementary | Broom & Adams, 2009[41] | Knowledge of oncologists on CAIM interventions; experiences of oncologists | 1. Varied reactions from oncologists when patients disclosed CAIM use ranging from negative or dismissive approaches to acceptance. | 1. Lack of policy and clinical practice guidelines in Australia for integrating CAIM into biomedical cancer | Not available | There were various consultant approaches to CAIM, and risk was consistently |

| | | | | | | |
|---|---|---|---|---|---|---|
| and Alternative Medicine With Their Patients | | counselling cancer patients on CAIM and the approaches used in these encounters. | 2. Most oncologists were skeptical towards CAIM use and desired to limit use due to the perceived risk of harm, but also worried they would alienate patients upon dissuasion.<br>3. Risks were a prevalent topic during patient-physician discussions on CAIM. However, oncologists disagreed on their duty to warn patients of CAIM risks. Some oncologists believed it a responsibility to protect their patients, while others believed that, due to the minimal evidence on CAIM risks, advice should be withheld.<br>4. Oncologists' lack of knowledge of CAM interventions and their risks, coupled with resistance amongst physicians to non-biomedical educators, presented ethical concerns when counselling patients on CAIM use. | care.<br>2. Lack of education on CAIM is a challenge. | | deployed to tackle CAIM issues and potentially direct patient behaviour. CAIM education was a crucial issue for oncologists. Based on the findings, major clinical practice and work concerns in CAIM sociology were discussed. |
| Traditional Medicines, Collective Negotiation, and Representations of Risk in Indian Cancer Care | Broom & Doron, 2012[42] | Experiences of oncologists in discussing TCAM with patients and their families | 1. The collectivist culture of India plays a large role in the communication. Communication of diagnosis and treatment often occurs between physicians and patients' families without direct input from patients. Physicians attempt to resist this dynamic and disclose cancer diagnosis to | 1. Lack of formal training as part of the Indian Biomedical Curriculum and advanced oncology training.<br>2. The complex mix of ideologies, beliefs, and religions influences patients' | Not available | This article revealed the difficulties in communication in relation to cancer's relative novelty, cultural practices around collective negotiation, and rhetorical practices evident in advising |

| | | | | | | |
|---|---|---|---|---|---|---|
| | | | patients directly<br>2. Tensions arose with TCAM disclosure: Oncologists faced difficulties getting patients to disclose their use of TCAMs and further dissuade them from using them. Physicians' negative attitudes towards TCAMs discourage patients from disclosure.<br>3. Discussions of risk were central to clinical-physician communication on TCAMs; physicians dissuaded patients from TCAM use due to concerns about the impurity and pollution of TCAM remedies, which may lead to toxicity. However, this was deemed ironic as conventional treatments such as chemotherapy can also confer toxicity, so it became a matter of who can deliver those toxins to patients. | and clinicians' understanding of health and physician roles.<br>3. The varying approaches to healthcare, such as individualistic and collectivist, also pose a challenge for patient-physician communication.<br>4. Lack of cancer literacy amongst patients, exacerbated by a lack of health education services, was a barrier during patient-physician discussions of TCAM. | | on risk revolving TCAM. The article discovered that as cancer becomes a major burden in India, research studying various forms of expertise, political representation, and the nexus between traditional beliefs and techno-scientific development is urgently needed. |
| Bridging the Gap Between Attitudes and Action: A Qualitative Exploration of Clinician and Exercise Professional's Perceptions to Increase | Caperchione et al., 2022[43] | Factors impacting implementation of exercise communication and referral; integrated clinical approaches to exercise communication and referral in cancer care. | 1. Oncologists were not comfortable discussing exercise as they lacked knowledge in general, and for cancer care specifically.<br>2. There is a need to support more education and training of oncologists with promoting exercise.<br>3. Oncologists were concerned about financial barriers for | 1. Lack of familiarity, knowledge and training on exercise generally, and in the context of cancer care.<br>2. Lack of direct referral processes to exercise program is frustrating for | 1. Limited generalizability due to regionality of participants.<br>2. Not all care providers (e.g., surgical oncologists) were represented.<br>3. Self-selection of participants that likely | Barriers to exercise prescription and referral include lack of knowledge, training, and funding. Based on these findings, a pragmatic model is provided to guide implementation-referral, inclusive of |

| | | | | | | |
|---|---|---|---|---|---|---|
| Opportunities for Exercise Counselling and Referral in Cancer Care | | | patients if they were to recommend exercise.<br>4. The lack of direct referral processes to exercise programs is frustrating, impedes care, and confuses patients.<br>5. When communicating to patients, it was important to understand when the right time is to mention exercise, and to provide simple resources to avoid information overload. | physicians and patients. | perceive exercise as important. | oncologist-initiated communication exchange, relevant resources, and access to exercise professionals with cancer expertise. |
| Changing Physicians' Attitudes Toward Self-help Groups: An Educational Intervention | Carroll et al., 2000[44] | Changes in physicians' attitudes (positive or negative) after exposure to the educational package. | 1. Educational article improved physicians' sense of importance of initiating discussions with patients about self-help groups.<br>2. After education, some physicians changed their attitudes and planned to change practices to explore the potential benefits of self-help groups for their cancer patients.<br>3. Physicians believe it is important to be open to multidisciplinary care and appreciate the patient perspective, as doctors can't address the depth and range of many issues.<br>4. A small number of physicians still hold reservations about self-help groups and are concerned with misinformation and encouragement of unproven therapies. | Not available | 1. Response rate: only 41% of the original sample completed the study. 2. Time span: unclear whether changes in attitudes persisted as time passed or whether it translated into changes in practice.<br>3. It is possible a higher proportion of participants favourably disposed to self-help groups were included. | Having family physicians review education material specific to their concerns about self-help groups can change their attitudes towards cancer self-help groups. Further study is needed to see whether this method will translate into changing practice patterns. |

| | | | | | | |
|---|---|---|---|---|---|---|
| Oncologists' Experiences of Discussing Complementary and Alternative Treatment Options With Their Cancer Patients. A Qualitative Analysis | Corina et al., 2016[45] | Considerations made by oncologists when discussing CAIM with patients; experiences with such discussions during and after the conventional oncological treatment phase; challenges in discussing CAIM; role oncologists adopt during CAIM discussions. | 1. Physicians used language that suggests "otherness" regarding CAIM remedies and CAM belonging to "another world."<br>2. During interviews, the boundaries between CAIM and CONV became blurred, with some oncologists showing openness and acceptance of CAIM.<br>3. Physicians described 'talking about CAIM' and providing advice on it as services they offer patients.<br>4. Although some oncologists were skeptical about CAIM, they attempted to be open-minded to foster trust with patients during clinical visits.<br>5. Counselling on CAIM includes warning patients of harm to protect them from negative health and financial outcomes, with many physicians emphasizing the lack of scientific research on CAIM efficacy and safety to dissuade patients. This may be discouraging to patients if it is the only content of the discussion. | 1. Lack of formal training and established standards for discussing CAIM with patients.<br>2. Lack of knowledge of the subject, and weak results from scientific studies make it difficult to justify certain recommendations on CAIM.<br>3. Lack of clear definition of CAIM. | 1. Low number of telephone interviews initially conducted; the convenience sampling technique may have resulted in a homogenous set of interviewees. | Training and educational resources should be made available to cancer specialists to expand their understanding of CAIM's potential benefits and harms. This would help to reduce the perceived disparities between CAIM and CONV among experienced oncologists. Furthermore, education in CAIM should be part of formal medical training as this would prevent students from feeling that CAIM belongs to another world. |
| Complementary Medicine for Cancer Patients in General Practice: Qualitative | Dahlhaus et al., 2015[46] | Reactions and experiences of GPs when patients show interest in CAIM; the relation between | 1. GPs described general practice as providing 'spoken medicine' in contrast to specialist care, which is why they felt their advice on CAIM | 1. Lack of CAIM-specific training for GPs.<br>2. Negative attitudes and lack of open- | 1. Small sample size.<br>2. Deliberate use of provocative language, such as the statement 'CAIM use out of a | Although oncology GPs view themselves as key information sources on CAIM for their patients, they |

| | | | | | | |
|---|---|---|---|---|---|---|
| Interviews with German General Practitioners | | these reactions and their knowledge of CAIM. | was necessary.<br>2. Many GPs felt responsible for preventing cancer patients from using non-evidence-based CAIM therapies.<br>3. GPs reported a lack of institutional CAIM education programs, and the poorly researched and large CAIM market made it challenging for individuals who did not welcome CAIM to gain more insight.<br>4. GPs must be open-minded to CAIM in order to self-directed learning. Otherwise, they would make recommendations during patient visits based on personal experiences from daily practice.<br>5. When discussing CAIM with patients, many GPs mainly reacted to patients' requests and information needs but did not proactively advise cancer patients on CAM or even ask about CAIM use.<br>6. Due to a lack of knowledge of CAIM, some GPs make recommendations to patients during visits based on personal experiences from daily practice.<br>7. Conflicting views existed on the role of GPs in encouraging CAIM use: some affirm the value of CAIM to every patient that uses it regardless of | mindedness on the part of physicians are barrier to seeking knowledge on CAIM. | feeling of powerlessness.'<br>3. Did not provide physicians with a definition of CAIM. | are uncertain and unfamiliar with the topic. GPs would benefit from CAIM training that allows them to discuss this topic with their patients proactively, openly and honestly. |

| | | | | | | |
|---|---|---|---|---|---|---|
| | | | whether they are convinced of its efficacy due to feelings of powerlessness and in attempts to foster positivity, trust, and hope. Others are hesitant to do so due to safety concerns stemming from a lack of evidence thus they provide support, empathy and care by maintaining honest communication<br>8. GPs are often the first individuals with whom patients share their CAIM use or seek CAIM advice. | | | |
| A Qualitative Study of Patient and Healthcare Provider Perspectives on Building Multiphasic Exercise Prehabilitation into the Surgical Care Pathway for Head and Neck Cancer | Daun et al., 2022[47] | HCPs' and patients' perceptions of incorporating multiphasic exercise pre-habilitation into the HNC surgical care timeline; physicians' level of ease in implementing the additional assessment to usual care; clinical logistics and benefits of the exercise rehabilitation. | 1. Ongoing, clear communication allowed for the development of new and effective workflow strategies for HCPs.<br>2. The exercise program allowed patients to communicate their experiences and fostered a sense of hope.<br>3. HCPs noted improvements in patient recovery, emotional and functional well-being, hospital length of stay, and wound recovery.<br>4. HCPs believed that exercise specialists should be involved in the care pathway.<br>5. Exercise should be comprehensive and tailored to each patient.<br>6. Key areas for implementation | 1. Lack of administrative and policy support for the integration of exercise programs in cancer care.<br>2. Time-sensitivity of the cancer condition made exercise implementation challenging.<br>3. The exercise program added additional documentation to the standard clinical workflow. | 1. Missing perspectives of other key stakeholders (e.g., administrative, policy).<br>2. Small sample size.<br>3. Sample included mainly self-reported White males.<br>4. Potential researcher bias, as the lead researcher is White, cis-female, and has never been diagnosed with cancer. | Findings informed the implementation of an exercise program in HNC surgical patient care. Future implementation must prioritize key stakeholder perspectives and be influential across multiple levels to support behaviour change. |

| | | | | | | |
|---|---|---|---|---|---|---|
| | | | included more education for patients and HCPs, a cultural shift in clinical care for cancer, and a collaborative team approach with engagement with the surgical team. | | | |
| The Challenge of Timing: A Qualitative Study on Clinician and Patient Perspectives About Implementing Exercise-Based Rehabilitation in an Acute Cancer Treatment Setting | Dennett et al., 2020[48] | Barriers and facilitators to exercise-based rehabilitation during treatment; considerations to provide rehabilitation while undergoing treatment; what HCPs believe is the most important element of rehabilitation for cancer patients. | 1. It is beneficial to implement rehabilitation early after diagnosis but the timing should be individualized depending on patient needs.<br>2. Exercise rehabilitation was seen as positive and important in cancer care, including to potentially improve tolerance to anti-cancer therapies and influence disease outcomes.<br>3. Clinicians reported low awareness and lacked confidence on what to advocate for with regards to rehabilitation exercise, accordingly offering vague advice.<br>4. There was poor knowledge about available resources on rehabilitation.<br>5. Convenience factors of the rehabilitation program such as duration, time, and location, were critical to facilitating referrals.<br>6. it was important for clinicians to refer patients to staff trained in oncology rehabilitation, | 1. Lack of policies supporting the integration of exercise programs in cancer care.<br>2. Lack of resources and services to refer patients to deterred physicians from counselling patients on exercise and rehabilitation programs. | 1. Lack of generalizability to all cancer survivors.<br>2. Potential sample bias as cancer survivors with an interest in exercise may have been more inclined to participate. | Exercise-based rehabilitation is necessary but challenging. Attitudes, knowledge, access to resources, and convenience of the program are important to consider. |

| | | | | | | |
|---|---|---|---|---|---|---|
| | | | equipped with skills to manage low motivation of patients. | | | |
| A Comparison of Physician and Patient Perspectives on Unconventional Cancer Therapies / Physician Perspectives on Unconventional Cancer Therapies | Gray et al., 1997; Gray et al., 1998[49,50] | Experiences with unconventional therapies; patient-physician communication issues involving unconventional therapies. | 1. Physicians believed it was important to have information available on unconventional therapies even if they were not always keen on them, because of the importance of keeping patients informed and respecting their autonomy.<br>2. Many physicians believed their role was as a gatekeeper and interpreter of information, while only some believed they should directly facilitate patient access to information.<br>3. Most physicians supported patients' intent in using unconventional therapies, but this was limited to therapies complementary to conventional medicine and safe for patients physically and financially.<br>4. The doctor-patient relationship was improved when physicians showed interest in patients' choices, even in cases where the physician disagreed with their choices.<br>5. Most physicians were not interested in collaborating with unconventional practitioners because they either wanted nothing to do with them or they believed the practices were | 1. Systemic barriers such as time restrictions in clinical settings hinder communication.<br>2. Physicians needed more information regarding unconventional treatments and noted that undergraduate medical education overlooks unconventional therapies.<br>3. Many physicians were not inclined to initiate conversations on CAIM due to the fear of being perceived as uninformed or dismissive. | 1. Some physicians were part of the research team which may limit generalizability and increase risk of sample bias.<br>2. Lack of diversity in the physician sample.<br>2. Physicians who bought "A Guide to Unconventional Cancer Therapies" may have been more accepting towards unconventional approaches which increases the risk of biased results. | Cancer specialists, family physicians, and other health professionals should look for ways to meet the needs of their patients interested in unconventional approaches to preserve and improve provider-patient relationships. Suggestions for future research, as well as educational and policy strategies were addressed. |

| | | | |
|---|---|---|---|
| | | | mutually exclusive, although they were willing to work together in research contexts to evaluate the effectiveness of unconventional therapies.<br>6. Physicians reported that patients interested in unconventional therapies fall into one of the following categories: the intelligent, the desperate, the gullible, the ignorant, the anti-establishment, the ethnic, and the very sick.<br>7. Physicians believed that unconventional therapies were popular among patients because of appealing marketing strategies, the sense of personal control they provide, mistrust of conventional medicine, and the support they receive from unconventional therapy providers.<br>8. Physicians pointed the blame towards patients regarding poor physician-patient communication due to patients' unrealistic expectations, intolerant towards reality, hostility towards or denial of bad news, disorganization in seeking information, and non-disclosure of unconventional therapy use.<br>9. Many physicians recognized |

| | | | | | | |
|---|---|---|---|---|---|---|
| | | | the lack of credibility of unconventional therapies' usefulness and reported a distrust towards unconventional therapy practitioners who often seduce patients into using unproven interventions.<br>10. Physicians believed their patients' choices to pursue unconventional therapies are illegitimate from a medical perspective and expressed concerns about dangers. | | | |
| What Hinders Healthcare Professionals in Promoting Physical Activity Towards Cancer Patients? The Influencing Role of Healthcare Professionals' Concerns, Perceived Patient Characteristics and Perceived Structural Factors? | Haussmann et al., 2018[51] | HCPs' knowledge; perceived advantages, and disadvantages of PA; facilitators and barriers for PA promotion. | 1. HCPs had a positive attitude towards PA but were concerned about physical overexertion, which led them to recommend modified or adjusted exercise if needed.<br>2. HCPs were concerned that encouraging PA would result in unrealistic expectations that could lead to psychological stress from missing exercise.<br>3. HCPs were more likely to promote PA if they believed patients to have an interest in PA by considering patient's past PA experience.<br>4. HCPs were hesitant to discuss PA out of fear that they would jeopardise a good relationship with patients, and that patients may take offence or block out advice relating to PA.<br>5. Some HCPs preferred | 1. There was limited time during clinic visits which pushed PA promotion to the bottom of the priority list.<br>2. HCPs reported a lack of knowledge on whether to or how to promote PA, and wanted more educational information for themselves and their patients. | 1. Study sample limited to the German federal state of Baden-Württemberg and may lack generalizability to other countries.<br>2. Low participation rate for specialized physicians, leading to a selective study population.<br>3. The focus on only 3 factors may have prevented the generation of other relevant findings. | The findings enabled a thorough examination of HCPs' considerations and influential factors of PA promotion. HCPs were primarily concerned with a patient's physical condition and the risk of overexertion. The study also pointed to structural factors that hindered HCPs in promoting PA for patients with cancer. |

| | | | | | | |
|---|---|---|---|---|---|---|
| | | | referring patients to in-house structures in inpatient settings such as in-house physiotherapy as it would guarantee that patients receive a PA consultation.<br>6. Physicians expressed the importance of addressing PA earlier in the treatment stage as it would lead to better acceptance from patients. | | | |
| On Caring and Sharing–Addressing Psychological, Biographical, and Spiritual Aspects in Integrative Cancer Care: A Qualitative Interview Study on Physicians' Perspectives / The Subjective Dimension of Integrative Cancer Care: A Qualitative Study Exploring the Perspectives, Themes, and Observations of Experienced Doctors from the Area of | Kienle et al 2018a; Kienle et al 2018b[52,53] | Physicians' perceptions; physicians' treatment goals; physicians' procedures and treatment decisions; physicians' observations regarding subjective and emotional dimensions of cancer diseases and cancer care including psychological and spiritual consideration; safety factors; new physician insights. | 1.When assessing patients, doctors took a patient-centered, multimodal treatment approach that employed conventional and complementary holistic concepts.<br>2. Following MT use, some physicians noted physical health improvements in patients, including brighter skin, appearing less tired, increase in appetites and weight, pain reduction, reduced susceptibility to colds and flus, and improved sleep quality.<br>3. MT use improved tolerance to anti-cancer therapies, including reduced side effects and better recovery.<br>4. MT use provided patients with psychological support, including supporting autonomy, coping, acceptance of cancer, hope, peace, self-confidence, | Not available | 1. Physicians' views were only described and not directly observed.<br>2. Results might be biased towards cases of patient satisfaction rather than those who were disappointed.<br>3. Purposefully recruiting an experienced group of physicians from German-speaking countries may limit generalizability of findings to the average caregiver.<br>4. Limited number of women in the sample.<br>5. Physicians' positive attitudes may have enhanced patient support that was attributed to MT use.<br>6. Integrative treatment | Physicians reported a range of positive observations from their patients' use of MT, including improved strength, warmth, improved psychological states, and resulting in a positive outlook on life and their future. Future studies should explore how to sufficiently meet the needs of patients and the influence of interventions such as MT on patient. |

| | | | | | | |
|---|---|---|---|---|---|---|
| Anthroposophic Medicine | | | initiative, and spirituality/connection with oneself.<br>5. Physicians emphasized the importance of respecting patients' autonomy and informed choice-making.<br>6. Physicians' attitudes about spirituality may influence how they support patients engaged in spiritual practices. | | settings impede the possibility of causal attributions to interventions. | |
| German Physicians' Perceptions and Views on Complementary Medicine in Pediatric Oncology: A Qualitative Study | Klatt et al., 2023[54] | German POs' understanding of CAIM; experiences, challenges, and strategies for discussing CAIMs. | 1. POs perceived "complementary" positively, but "alternative" medicine as concerning with regard to efficacy and the potential of harm in replacing traditional treatments.<br>2. POs believe that CAIM may improve disease symptoms but not actually treat the cancer itself.<br>3. POs recognized that CAIM use provided patients with a sense of empowerment and control.<br>4. POs considered several possible types of harm can be conferred from CAIM use.<br>5. POs viewed being open to CAIM discussions as professional behavior to prevent harm and some advocated for a proactive approach to raise the topic.<br>6. POs were willing to search for | 1. It is difficult to decide how or when to intervene regarding the possible negative effects of CAM.<br>2. Barriers to discussing CAM included uncertainty of the evidence and challenges in gaining knowledge on the efficacy, risks, costs, and feasibility of CAIM.<br>3. There was a lack of time in daily routines to discuss CAIM. | 1. Lack of generalizability to other clinicians.<br>2. The study may have attracted only POs already interested in CAIM.<br>3. Snowball sampling could have limited diversity of participant views on CAIM.<br>4. Findings require future triangulation with views of other stakeholders (other practitioners, families of patients etc.). | The interdisciplinary interpretation of findings by experts from various health disciplines suggests that there is a need to establish a consensus on the minimal professional standards on addressing CAIM in pediatric oncology settings. |

| | | | | | | |
|---|---|---|---|---|---|---|
| | | | information on CAIM and often relied on professional networks of CAIM practitioners.<br>7. POs believed they had an obligation to intervene if CAIM posed physical harm or high financial burden for patients. | | | |
| Lifestyle Advice to Cancer Survivors: A Qualitative Study on the Perspectives of Health Professionals | Koutoukidis et al., 2017[69] | HPs' awareness of survivorship lifestyle guidelines; barriers to provision of lifestyle advice; views on various formats for lifestyle interventions. | 1. HPs believed lifestyle behaviour change was particularly challenging for survivors due to ingrained life long habits and low mood and distress.<br>2. Sensitive topics such as alcohol and tobacco use were not addressed unless initiated by patients.<br>3. Advice was tailored to each patient, and frailty or physical limitations significantly hindered the inclination to provide advice.<br>4. HPs had concerns with straining the patient-physician relationship upon advising patients to do PA.<br>5. HPs regarded lifestyle changes and PA to be of lower priority to cancer survivors.<br>6. HPs varied in their beliefs of how patients will perceive lifestyle advice.<br>7. HPs perceived their own lack of adherence to lifestyle guidelines as both a barrier and facilitator to counselling | 1. HPs noted that survivors faced practical socioeconomic barriers (e.g., affordability of healthy diet, gym membership).<br>2. Lack of social support and cultural variation made promotion of health behaviours difficult.<br>3. Limited time during clinic visits was a challenge.<br>4. HPs had a lack of awareness and knowledge on lifestyle guidelines which hindered the confidence of HPs to counsel on such changes. | 1. Lack of generalizability to all oncology HPs.<br>2. Potential selection bias as participants may have been generally more interested in lifestyle topics. | Some but not all HPs specialising in breast, prostate and colorectal cancers provide lifestyle advice to cancer survivors post-treatment. HPs who provide advice rely on guidelines for the general population with attempts to tailor it to individuals. More education and training programs are needed to increase the effectiveness of counselling. |

| | | | | | | |
|---|---|---|---|---|---|---|
| | | | patients.<br>8. HPs noted the importance of tailoring plans for individuals.<br>9. HPs were conscious to not overwhelm patients with multiple changes at once and believed that framing the advice as part of patients' treatment was effective. | | | |
| Academic Doctors' Views of Complementary and Alternative Medicine (CAM) and Its Role Within the NHS: An Exploratory Qualitative Study | Maha & Shaw, 2007[55] | Background/training on CAM; views on CAIM; extent of professional interest in CAIM; views of patient demand for CAIM; views of own knowledge on CAIM; views of usefulness of CAIM and for what conditions (if any); views of appropriate settings in which to provide CAIM. | 1. Perspectives on CAIM varied, including positive (from physicians who underwent additional training and/or provide CAIM), negative/skeptical, and undecided.<br>2. Physicians who offered CAIM believed it allowed them to practice medicine more holistically, allowed patients to feel heard, and enabled them to address beneath-surface symptoms.<br>3. Physicians with negative views distinguished between therapies that claimed to 'cure' patients and those that offered supportive care, and favoured the latter.<br>4.Neutral physicians said they lacked knowledge on CAIM to make a judgement on the efficacy.<br>5. Physicians rarely initiated conversations with patients on CAIM as they felt it was not a | 1. Costs were a barrier to discussing CAIM as these patient-physician encounters tended to be longer and more costly than conventional consultations.<br>2. Physicians reported insufficient CAIM knowledge for them to make informed judgements on the effectiveness.<br>3. Most physicians lacked formal training in CAIM and had insufficient knowledge. | 1. Small sample size.<br>2. Did not assess the credibility of themes and their interpretation at earlier stages in the analysis process.<br>3. Lack of generalizability to non-academic doctors and doctors across diverse settings.<br>4. Potential underrepresentation of middle-ground views on CAIM. | Providers that don't practice CAIM were skeptical or uncertain about the value of CAIM, which was a barrier to CAIM integration. However, it is important that physicians facilitate an atmosphere of openness regarding consultations so patients are able to discuss CAIM. Open physician-patient communication can allow for concerns to be addressed, and enhance physician knowledge on what patients are using. |

| | | | | | | |
|---|---|---|---|---|---|---|
| | | | priority especially in cases when the scientific evidence was weak.<br>6. Most physicians only consulted patients on CAIM to better understand their usage as opposed to agreeing with CAIM use.<br>7. Patient choice influenced whether physicians discussed or suggested CAIM use.<br>8. Physicians acknowledged that their skepticism towards CAIM may be a barrier to patients when considering CAIM use disclosure or requesting a referral to a CAIM practitioner.<br>9. Most physicians wanted further research on CAIM to establish specific modalities and 'gold standards.'<br>10. Physicians varied on their views of integrating CAIM within the NHS. | | | |
| Yoga in Adult Cancer: A Pilot Survey of Attitudes and Beliefs Among Oncologists | McCall & Heneghan, 2015[56] | Oncologists' open remarks about yoga in cancer care<br>Note: extracted findings have been limited to qualitative (open-ended) data. | 1. Oncologists believed yoga was a gentle exercise worth exploring for symptom management for relaxation and stress management.<br>2. Barriers identified by oncologists include physical (ex. fatigue, pain, nausea, fear of infection), logistical (finances, transportation, scheduling), psychological (stress, anxiety, depression), and yoga-related | 1. Difficulty in recommending yoga due to limited scientific evidence and lack of knowledge on how and where to access classes. | 1. Risk of reporting bias.<br>2. Low response rate<br>3. Qualitative components limited to 3 open ended questions and one way communication. | Oncologists identified a lack of scientific evidence and other barriers to support the implementation of yoga for adult cancer patients. Oncologists would support conducting research to learn better about the |

| | | | | | | |
|---|---|---|---|---|---|---|
| | | | (lack of patient-perceived benefits or interest). | | | impact of yoga on cancer. |
| How is Complementary Medicine Discussed in Oncology? Observing Real-Life Communication Between Clinicians and Patients with Advanced Cancer | Mentink et al, 2022[57] | Physician-patient communication on CM, including who introduces the topic and when, what aspects are mentioned, and how attitudes are verbalized. | 1. When clinicians introduced CM into the conversation, it was mainly about nutrition and lifestyle. They were also encouraging about CM relating to nutrition of physical interventions, referencing scientific evidence on the topic.<br>2. Clinicians did not inquire about interest or use generally, and instead mentioned specific CM modalities directly.<br>3. Some clinicians disregarded CM statements made by patients and only a minority followed up with questions.<br>4. Clinicians replied to patients' questions about CM efficacy by stating that scientific evidence is limited or absent.<br>5. Clinicians were often the ones to suggest alternatives for the type of CM their patient was using.<br>6. Clinicians were highly unlikely to contact or refer patients to CM providers.<br>7. Clinicians emphasized the importance of patient autonomy and choice in terms of CM use.<br>8. Clinicians overall verbalized encouraging or neutral attitudes towards CAM. | Not reported. | 1. Assessment of validity and transferability of the observation scheme is needed.<br>2. Sharing of test results at follow up visits may have influenced the likelihood of discussions about CM to arising during consultations.<br>3. Lack of consensus on CM definition limits generalizability of findings. | HCPs are less likely to initiate CM discussions than patients. Ensuring routine discussions of CM use by patients during clinical encounters would prevent risks and maximize the benefits of CM use for patients with cancer. Future areas of investigation are the needs and expectations of clinicians and patients with cancer in regard to discussing CM use in daily oncology practice. |

| | | | | | | |
|---|---|---|---|---|---|---|
| | | | 9. Many clinicians suggested adding or changing conventional treatment rather than CMs. | | | |
| Oncologists’ and Naturopaths’ Nutrition Beliefs and Practices | Novak & Chapman, 2001[58] | Beliefs about the role of diet in breast cancer etiology; the relationship between those beliefs and scientific and other kinds of evidence; and the influence of those beliefs and the use of evidence on practitioners’ counseling practices. | 1. While oncologists acknowledged that diet affected health in general, they did not view it as specific to cancer prevention, prognosis or risk of recurrence.<br>2. Oncologists believe discussion and recommendations of diet are secondary priorities in clinical encounters.<br>3. Oncologists indicated that their guidance to patients primarily involves general advice on maintaining a healthy diet. | 1. Oncologists reported a lack of scientific evidence, or conflicting evidence, specifically randomized controlled trials, on the impact of diet on breast cancer prevention and treatment. | 1. Lack of generalizability due to the exploratory nature and small sample size.<br>2. Potential for bias from informants and during data analysis.<br>3. The results presentation may be biased towards what is more acceptable in a scientific context due to the author's affiliation with a conventional teaching and research institution. | There are fundamental philosophical and ideological differences between oncologists and naturopaths. Informing patients about these distinctions can empower them to make better-informed healthcare decisions. |
| Complementary Therapy Use by Cancer Patients: Physicians’ Perceptions, Attitudes, and Ideas | O'Beirne et al., 2004[59] | Perspectives on patients' CM use. | 1. Physicians found it difficult to define complementary therapies.<br>2. Physicians expressed the importance of holistic health but identified conflicts between this and evidence-based treatment.<br>3. There was a range of attitudes towards CM, from completely unsupportive, to those who actively recommended it.<br>4. Physicians believed patients used CM for reasons such as lack of benefit from | 1. Lack of education or knowledge on CM, which presented as a communication barrier with patients.<br>2. Lack of standardized definition for complementary therapies.<br>3. Lack of scientific evidence for CM efficacy. | 1. Small sample size.<br>2. Men are overrepresented in the sample. | There is a need for educational programs or future research on CM during clinical encounters, including physicians’ role in patients’ decision making about complementary therapies, ways to improve patient-physician communication about |

| | | | | | | |
|---|---|---|---|---|---|---|
| | | | conventional therapies, to manage side effects, please family members, gain social support, deal with guilt of their poor lifestyle, desire for hope and control, and the perception that CM was less toxic because it was "natural."<br>5. Physicians believed it was important for discuss this with patients for better communication.<br>6. Physicians recognized that most of their patients do not disclose CM use due to fear of rejection and embarrassment, poor communication experiences with physicians, the belief that family physicians have little knowledge of CM, and fear of conflicting with the physicians' belief systems.<br>7. Physicians identified their role as offering support, education, protection from harm, and providing complementary treatments.<br>8. Physicians were concerned that showing support to discuss CM might be interpreted as them supporting the therapy itself.<br>9. Physicians were concerned with the lack of accountability of CM providers, and financial and psychological harms (i.e., | | | complementary therapies, ways to communicate bad news so that patients do not discourage patients from seeking conventional therapies, and the role and quality of scientific evidence on the use of complementary therapies. |

| | | | | | | |
|---|---|---|---|---|---|---|
| | | | false hope) associated with CM treatment. | | | |
| Perspectives of Pediatric Oncologists and Palliative Care Physicians on the Therapeutic Use of Cannabis in Children with Cancer | Oberoi et al., 2022[60] | Oncologists' level of education on medical cannabis; desired knowledge about medical cannabis; perspectives on the use of medical cannabis in pediatric oncology; concerns with using medical cannabis; attitudes towards future research. | 1. Physicians found that caregivers had a growing interest in cannabis use, due to strong beliefs in its positive effect.<br>2. Physicians also expressed the importance of patient engagement, shared-decision making, and using a non-judgmental approach.<br>3. Physicians were concerned about the possible physical harms to children and the financial burden of cannabis use. | 1. Lack of institutional and organizational policies on cannabis use for children with cancer.<br>2. Lack of robust research findings regarding cannabis' effect in cancer care.<br>3. Physicians identified the need to combat inaccurate information from social media and web sources. | 1. Results lacked perspectives on the use of THC and CBD.<br>2. Demographic information of participants was not collected.<br>3. The number of patients using cannabis could have been overestimated or underestimated.<br>4. Selection bias as physicians with no cannabis experience, or that do not believe it is relevant, could have opted out of participating, thereby limiting generalizability. | Most physicians recognize the potential of cannabis for symptom relief. More research is needed to facilitate informed decision-making in collaboration with pediatric oncology patients and their caregivers. |
| Convergent Priorities and Tensions: a Qualitative Study of the Integration of Complementary and Alternative Therapies with Conventional Cancer Treatment | River et al., 2018[61] | Oncologists' experiences and views of delivering integrative cancer care services. | 1. Oncologists valued a person-centred approach but were concerned with the evidence-base of CAIM.<br>2. Oncologists cautioned patients about certain CAIMs and made more modest recommendations.<br>3. Oncologists believed they should avoid "open support" for CAIM due to fears of CAIM safety.<br>4. Oncologists often found it hard to believe medical | 1. Cost to the interventions is a major issue. | 1. Small sample size limits generalizability to other cancer care settings. | Recognizing points of convergence and divergence between patients, nurses, and oncologists on CAIM integration could aid clinicians and service providers in navigating paths forward for integrative cancer services. |

| | | | | | | |
|---|---|---|---|---|---|---|
| | | | evidence citing the efficacy of CAIM.<br>5. Oncologists identified a tension between patient needs and limited resources, and the need for more evidence to justify spending public dollars on CAIM. | | | |
| Mind the Gap! Lay and Medical Perceptions of Risks Associated with the Use of Alternative Treatment and Conventional Medicine | Salamonsen, 2015[62] | Physicians' experiences with patients' CAIM use, risk perceptions associated with conventional medicine and CAIM, and physician-patient communication. | 1. Physicians of different specialists had a common understanding of risks.<br>2. Oncologists had more concerns about CAIM than neurologists and GPs based on negative consequences of patient use observed in their oncological practice.<br>3. Physicians also trained in CAIM believed most were safe and beneficial. However, they believed that some CAIMs and unauthorized CAIM practitioners could represent risk to patients.<br>4. Some CAIM therapies (acupuncture) were perceived as uncomplicated, whereas others (St. John's wort) was perceived as risky.<br>5. Risk assessment and risk communication was heavily influenced by acuteness and prognoses of patient's disease.<br>6. All physicians expressed the need for scientific risk evaluations of specific CAIM | 1. Several physicians observed negative interactions between CAIM therapies and conventional treatment, leading to death of patients. This was concerning, and "scary."<br>2. The Norwegian Pharmacovigilance Advisory Board is unable to address negative consequences of CAIM and conventional treatment interactions as the CAIM treatment is not registered as a drug.<br>3. Physicians felt frustrated and insecure when seriously ill patients did not trust conventional medicine, and chose to delay or deny it in | 1. Results cannot be generalized to populations of cancer or MS patients or transferred to other diagnostic groups. | There are fundamental gaps in risk perceptions associated with the use of conventional medicine and CAIM among MS and cancer patients and their doctors. These differences influenced risk communication and patients' decision-making. Physicians believed in the safety of conventional medicine, but believed CAIM was potentially risky, which is contrary to patients. |

| | | | | | | |
|---|---|---|---|---|---|---|
| | | | therapies, and the need for more knowledge about communication with users of CAIM. | favour of conventional treatment. | | |
| ‘They Don’t Ask Me So I Don’t Tell Them’: Patient-Clinician Communication About Traditional, Complementary, and Alternative Medicine | Shelley et al., 2009[63] | Factors influencing attitudes toward TM/CAIM; experience in communicating with patients about TM/CAIM. | 1. Some clinicians believed TM/CAIM discussions were important for respecting patient autonomy and culture, but remained cautious regarding the efficacy, cost, and safety.<br>2. Clinicians believed it was important not to be dismissive of TM/CAIM use, and that language was important to establish a non-judgemental environment.<br>3. Some clinicians viewed themselves as scientific experts and felt obligated to "do no harm," leading them to warn patients of their concerns with TM/CAIM practices.<br>4. Clinicians rarely initiated conversations as they did not perceive a high level of CAIM use amongst their patients.<br>5. Some clinicians, concerned about the safety and efficacy, took an assertive approach to dissuading patients from TM/CAIM use. | 1. Many clinicians believed their knowledge of TM/CAIM was insufficient, which was a barrier to initiating conversations on TM/CAIM use.<br>2. The time constraints associated with brief clinical visits with patients limited when or how clinicians discussed TM/CAIM. | 1. Study took place in Hispanic and Native American communities where TM/CAIM is valued, which could limit generalizability.<br>2. Data was collected only from primary care clinics, and may not represent perspectives of people outside of primary care. | Communication barriers limit the extent to which patients and clinicians discuss TM/CAIM. Clinicians should change their communication dynamic to overcome barriers in discussing TM/CAIM use. |
| Attitudes Toward Complementary and Alternative Medicine Amongst | Siegel et al., 2016[64] | Views of HCPs on CAIM in cancer care; potential for CAM integration in oncology care. | 1. Some physicians showed support for CAM integration in cases where biomedical interventions were hopeless, or not sufficiently addressed by | 1. The integration of CAIM could be an additional resource burden with resource allocation/resource | 1. Data was collected in 2011 and published in 2016, and thus may be outdated.<br>2. Participants were | Exploring the attitudes of oncology health professionals in Brazil toward CAIM |

| | | | | | | |
|---|---|---|---|---|---|---|
| Oncology Professionals in Brazil | | | conventional medicine (e.g., palliative care, pain relief).<br>2. Most physicians were benevolent on the topic of CAIM and supportive of selectively integrating CAIM into cancer care, with significant reservations.<br>3. Some physicians expressed negative beliefs towards CAIM, arguing that strong scientific evidence is needed to justify the integration of CAIM and the use of resources on it. | burden in an already overburdened field of cancer services.<br>2. Lack of knowledge or training on CAIM.<br>3. Lack of scientific evidence on CAIM. | from one cancer center, reducing the finding's generalizability. | remains a crucial area for research. These findings elucidate the perspectives held by health professionals on CAIM. As observed in global research, attitudes of health professionals toward CAIM impact patient utilization, as well as their interactions with conventional practitioners. Differences in views between patients and oncologist professionals can lead to communication challenges and potential breakdown of conventional oncological care. |
| Considerations in Developing and Delivering a Non-Pharmacological Intervention for Symptom Management in Lung Cancer: The Views of | Wagland et al., 2012[65] | HCPs views on delivery; uptake; future implementation and cost-effectiveness of an NPI. | 1. Non-pharmacological managements are often under-used and unfamiliar to oncologists.<br>2. All HCPs agreed that NPIs are essential to offer psychological support and a sense of control for patients, and should be integrated into treatment.<br>3. HCPs recognized the need for | 1. Logistical difficulties exist when administering NPIs in a hospital setting such as securing an appropriate room and scheduling protected nursing time.<br>2. HCPs also identified that patients | 1. Small sample sizes limits the study power and generalizability. | HCPs believe NPIs should be personalized to patients' individual needs and made available for patients when they are ready and receptive. NPIs are most effective if delivered to patients |

| | | | | | | |
|---|---|---|---|---|---|---|
| Healthcare Professionals | | | tailored patient education due to highly individualized needs.<br>4. Patients were more likely to accept NPIs that were tailored, recommended by HCPs, and the provider had sufficient training and belief in the intervention.<br>5. Most HCPs agreed that NPI use should be introduced once patients have completed their first cycle of treatment, not at initial diagnosis.<br>6. HCPs expressed that patients with lung cancer may be more dismissive of NPIs than other cancer groups due to SES and demographic factors.<br>7. NPIs should be delivered in a relaxing environment, such as a patient's home, over a hospital setting. | sometimes face issues with lack of affordable transportation when requiring repeated visits to an outpatient clinic. | | individually instead of in group settings, and outside of an acute medical facility. HCPs believed that a trained provider, and not a volunteer or lay person, should deliver these interventions. |
| ‘Probably Better Than Any Medication We Can Give You’ | Waterland et al., 2020[66] | GPs' views and experiences of providing nutrition and exercise advice. | 1. All GPs recognized the importance of exercise and nutrition for cancer patients and their role in ongoing communication on this.<br>2. Patients rarely attend clinical visits with exercise and nutrition on their agenda.<br>3. GPs noted that patients' sources of information were media and social networks.<br>4. GPs emphasized the importance of a tactful approach when providing recommendations, considering | 1. Lack of resources and programs supporting GPs in providing this advice.<br>2. A lack of knowledge on nutrition and exercise, which impacted their confidence in advising patients.<br>3. Limited time during consultations was a barrier to discussing nutrition and exercise. | 1. Potential for volunteer bias since GPs expressed interest in participating in the study. | This study offers evidence of GPs expressing a keen interest in actively supporting the healthy lifestyle habits of their cancer patients. It suggests potential directions for future research and the development of resources in this area to better equip |

| | | | | | | |
|---|---|---|---|---|---|---|
| | | | the delicate circumstances of patients with cancer.<br>5. Some GPs had preference to refer patients to clinicians within their own clinic. | 4. GPs reported that the lack of access to patients during treatment, the lack of referral pathways, and funding limitations restricted patient access to affordable allied health services. | | GPs in providing guidance. |
| Communication About Complementary and Alternative Medicine When Patients Decline Conventional Cancer Treatment: Patients' and Physicians' Experiences | Wode et al., 2023[67] | Similarities and differences between patient and physician experiences on treatment choices and communication. | 1. Physicians acknowledged the importance of CAIM to combat cancer, strengthen health, and provide holistic care.<br>2. All physicians were generally positive towards mind-body practices, but not multivitamin/mineral supplement, especially during chemotherapy.<br>3. Most physicians spoke to patients about the medical risks and financial cost of declining recommended oncological treatment for CAIM.<br>4. Physicians believed that most patients had negative experiences such as lack of effects, or side effects.<br>5. Physicians felt frustrated by patients' comments that physicians are narrow-minded or servants of the pharmaceutical industry.<br>6. Some physicians declined or were reluctant to abide by patients' wants to monitor | 1. Very little understanding regarding CAM among physicians.<br>2. Limited time for CAIM related discussions between patients and physicians.<br>3. Lack of knowledge inhibited physician communication with patients when the topic was raised.<br>4. It was challenging to convey knowledge on the natural course of untreated cancer and its consequences, as it was risky to be overly assertive, but it was important to ensure patients understood the implications. | 1. Lack of generalizability due to limited selection and inclusion criteria.<br>2. Data saturation was reached with a small sample size, making finding less representable. | Findings reveal the difficulty of shared decision-making in cases where patients' and physicians' views on treatment decisions are different. This emphasizes a need to address the complexity of these situations, considering patients' values, and improving physicians' knowledge on CAIM. |

| | | | | | | |
|---|---|---|---|---|---|---|
| | | | CAIM results with unconventional lab tests and examinations.<br>7. Physicians emphasized the need to show interest in CAIM use to not discourage communication and foster mutual trust.<br>8. Some physicians wished for a team of healthcare professionals with competency in CAIM to better support patients. | | | |
| Medical cannabis: An Oxymoron? Physicians' Perceptions of Medical Cannabis | Zolotov et al., 2018[68] | Physicians' views and experiences with medical cannabis generally; attitudes on recommending cannabis to patients; normative beliefs on cannabis. | 1. Physicians' views of cannabis as a non-medicine are influenced by the biomedical model, evidence-based medicine, and the status of cannabis as an illicit drug.<br>2. Physicians expressed that cannabis may contain multiple compounds with varying potencies, and the lack of standardization influenced their categorization of cannabis as non-medicine.<br>3. Physicians were concerned about integrating cannabis in their practice due to working alone without administrative or security staff and accordingly, fearing aggressive encounters with patients.<br>4. Physicians also often viewed cannabis as medicine in palliative care settings where | 1. There is a lack of scientific evidence on safety and efficacy of cannabis.<br>2. Absence of training programs for medical cannabis impeded physician acceptance and prescription.<br>3. Cannabis has a pre-existing impression as an illegal drug that poses substantial harm, resulting in some physicians overlooking its therapeutic effects. | Not reported | Physicians do not have a consensus on whether cannabis is a medicine or not, which suggests potential obstacles to implementing medical cannabis policies. Understanding physicians' perspectives and varying levels of willingness to adopt such policies is crucial for advancing policy in this evolving domain. |

| | | | |
|---|---|---|---|
| | | | the goal is not cure, but relieving patients of suffering without attaining a cure. |
| **Abbreviations:** AM: Anthroposophic Medicine; AT: Alternative Therapy; CAIM: Complementary, Alternative, and Integrative Medicine; CM: Complementary Medicine; CONV: Conventional Medicine; CTM: Complementary and Traditional Medicine; GP: General Practitioner; HCP: Healthcare Professional; HNC: Head and Neck Cancer; HP: Health Professional; ICC: Integrative Cancer Care; MT: Mistletoe Therapy; NPI: Non-Pharmacological Intervention; OMT: Osteopathic Manipulative Treatment; PA: Physical Activity; PCC: Patient Centered Care; PCP: Primary Care Provider; PO: Pediatric Oncologist; PRO: Patient Reported Outcome; QSR: Qualitative Systematic Review; SCO: Supportive Care in Oncology; SEGT: Supportive Expressive Group Therapy; TCAM: Traditional, Complementary, and Alternative Medicine; USA: United States of America | | | |
